# Automated Deep Learning-Based Detection of Early Atherosclerotic Plaques in Carotid Ultrasound Imaging

**DOI:** 10.1101/2024.10.17.24315675

**Authors:** Murad Omarov, Lanyue Zhang, Saman Doroodgar Jorshery, Rainer Malik, Barnali Das, Tiffany R. Bellomo, Ulrich Mansmann, Martin J. Menten, Pradeep Natarajan, Martin Dichgans, Marianne Kalic, Vineet K. Raghu, Klaus Berger, Christopher D. Anderson, Marios K. Georgakis

## Abstract

**Background:** Carotid plaque presence is associated with cardiovascular risk, even among asymptomatic individuals. While deep learning has shown promise for carotid plaque phenotyping in patients with advanced atherosclerosis, its application in population-based settings of asymptomatic individuals remains unexplored.

**Methods:** We developed a YOLOv8-based model for plaque detection using carotid ultrasound images from 19,499 participants of the population-based UK Biobank (UKB) and fine-tuned it for external validation in the BiDirect study (N = 2,105). Cox regression was used to estimate the impact of plaque presence and count on major cardiovascular events. To explore the genetic architecture of carotid atherosclerosis, we conducted a genome-wide association study (GWAS) meta-analysis of the UKB and CHARGE cohorts. Mendelian randomization (MR) assessed the effect of genetic predisposition to vascular risk factors on carotid atherosclerosis.

**Results:** Our model demonstrated high performance with accuracy, sensitivity, and specificity exceeding 85%, enabling identification of carotid plaques in 45% of the UKB population (aged 47–83 years). In the external BiDirect cohort, a fine-tuned model achieved 86% accuracy, 78% sensitivity, and 90% specificity. Plaque presence and count were associated with risk of major adverse cardiovascular events (MACE) over a follow-up of up to seven years, improving risk reclassification beyond the Pooled Cohort Equations. A GWAS meta-analysis of carotid plaques uncovered two novel genomic loci, with downstream analyses implicating targets of investigational drugs in advanced clinical development. Observational and MR analyses showed associations between smoking, LDL cholesterol, hypertension, and odds of carotid atherosclerosis.

**Conclusions:** Our model offers a scalable solution for early carotid plaque detection, potentially enabling automated screening in asymptomatic individuals and improving plaque phenotyping in population-based cohorts. This approach could advance large-scale atherosclerosis research.

GRAPHICAL ABSTRACT.
ASCVD – Atherosclerotic Cardiovascular Disease, CVD – Cardiovascular disease, PCE – Pooled Cohort Equations, TP– true positive, FN – False Negative, FP – False Positive, TN – True Negative, GWAS – Genome-Wide Association Study.

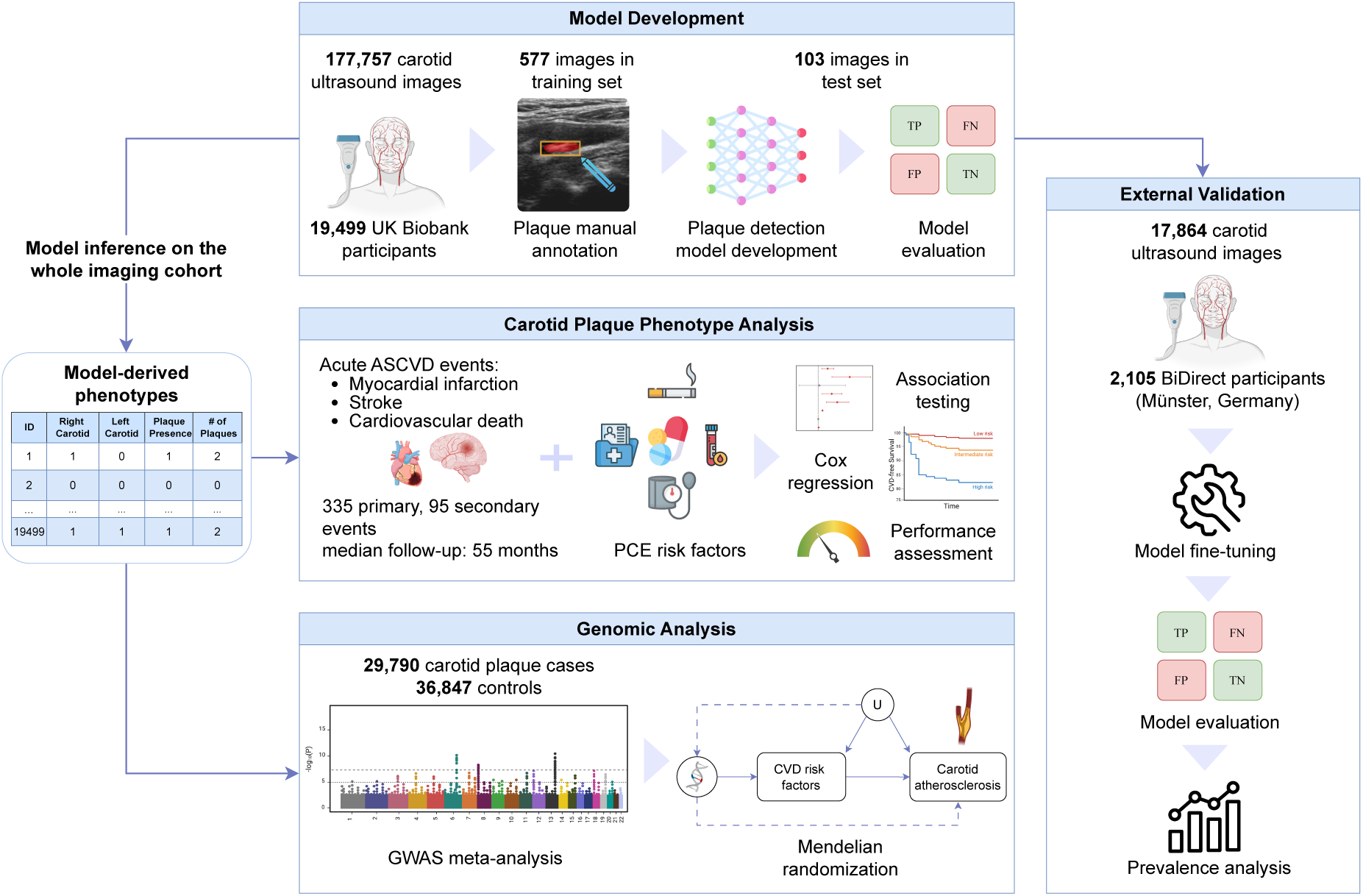

**CLINICAL PERSPECTIVE:** Carotid ultrasound is a well-established method for assessing subclinical atherosclerosis with potential to improve cardiovascular risk assessment in asymptomatic individuals. Deep learning could automate plaque screening and enable processing of large imaging datasets, reducing the need for manual annotation. Integrating such large-scale carotid ultrasound datasets with clinical, genetic, and other relevant data can advance cardiovascular research. Prior studies applying deep learning to carotid ultrasound have focused on technical tasks–plaque classification, segmentation, and characterization–in small sample sizes of patients with advanced atherosclerosis. However, they did not assess the potential of deep learning in detecting plaques in asymptomatic individuals at the population level.

We developed an efficient deep learning model for the automated detection and quantification of early carotid plaques in ultrasound imaging, primarily in asymptomatic individuals. The model demonstrated high accuracy and external validity across population-based cohort studies. Predicted plaque prevalence aligned with known cardiovascular risk factors. Importantly, predicted plaque presence and count were associated with future cardiovascular events and improved reclassification of asymptomatic individuals into clinically meaningful risk categories. Integrating our model predictions with genetic data identified two novel loci associated with carotid plaque presence—both previously linked to cardiovascular disease—highlighting the model’s potential for population-scale atherosclerosis research.

Our model provides a scalable solution for automated carotid plaque phenotyping in ultrasound images at the population level. These findings support its use for automated screening in asymptomatic individuals and for streamlining plaque phenotyping in large cohorts, thereby advancing research on subclinical atherosclerosis in the general population.

## Introduction

Atherosclerosis, characterized by lipid accumulation and plaque formation within arterial walls^1^, is the primary condition underlying cardiovascular disease (CVD), the leading cause of global mortality and morbidity^2,3^. Despite significant advancements in pharmacotherapies for lipid lowering and in the management of other vascular risk factors such as diabetes and hypertension, the alarmingly high and rising prevalence of CVD highlights the need for novel risk assessment and preventive strategies^4,5^. Atherosclerosis is a chronic disease, with subclinical atherosclerotic lesions developing silently over decades. Current CVD risk assessment tools used in clinical practice, including the Pooled Cohort Equations (PCE)^6^, Framingham Risk Score^7^, and Systematic Coronary Risk Evaluation (SCORE^8^ and SCORE2)^9,10^, rely on demographic, clinical, and biochemical factors but do not account for the presence of subclinical atherosclerosis^11,12^. Imaging studies that enable screening for subclinical atherosclerotic lesions in asymptomatic individuals suggest that atherosclerotic plaques are highly prevalent, even among individuals traditionally considered at low CVD risk^13–18^. Coronary artery calcium (CAC) scoring on computed tomography (CT) has gained traction for assessing subclinical atherosclerosis. However, it faces limitations as a screening tool due to ionizing radiation exposure and its relatively high cost for widespread application^19,20^.

In contrast, carotid ultrasound offers a non-invasive, radiation-free, and widely accessible modality for assessing subclinical atherosclerotic lesions^19,21^. While traditional assessment of carotid intima-media thickness (cIMT) does not reliably predict incident CVD risk^22^, the detection of carotid atherosclerotic plaques is associated with an increased risk of future events^21,23,24^. Despite promising results, it remains uncertain whether population screening for carotid plaques could reclassify asymptomatic individuals into higher-risk categories that justify the initiation of preventive pharmacotherapies^25^. Many published studies are constrained by relatively small sample sizes or insufficient follow-up^24,26–30^.

Several large-scale population-based cohorts have incorporated carotid ultrasound imaging into their data collection processes,^31–33^ but evaluating plaque presence across thousands of images remains a labor-intensive task. Recent advancements in deep learning have enhanced medical imaging analysis, enabling greater precision and the automation of processing large volumes of imaging data^34,35^. Although previous deep learning models for ultrasound images have shown potential in various tasks, including carotid wall and plaque segmentation^36–38^, these studies have focused on specific patient populations with advanced atherosclerotic disease, such as stroke patients or individuals with known carotid artery disease, which limits the generalizability of their findings to the broader population of asymptomatic individuals. Automating the detection of early carotid plaques in ultrasound images at a general population level could accelerate plaque screening in asymptomatic individuals and streamline plaque phenotyping in population-based cohorts. The integration of plaque phenotypes with other imaging modalities, as well as genetic, omics, and clinical data collected in the context of these studies would facilitate in-depth research into the biology of subclinical atherosclerosis, enabling explorations into the natural history of the disease and uncovering novel drug targets.

Here, we introduce a computer vision model designed to detect early atherosclerotic plaques in population-based settings. We utilized ultrasound images from 19,499 deeply phenotyped participants of the population-based UK Biobank (UKB). Our model demonstrated high performance in both detecting plaques and quantifying their counts, and showed external validity in a population-based cohort of 2,105 individuals in Germany (BiDirect). To demonstrate the potential of our model for population plaque screening and research into subclinical atherosclerosis, we leveraged its predictions to: (1) estimate age- and sex-specific prevalence rates of carotid plaques and compare them to those from other population-based cohorts applying manual plaque annotation; (2) confirm relationships with established vascular risk factors; (3) show associations of plaque presence and count with the risk of future CVD events; (4) assess whether plaque presence and count could improve CVD risk prediction; and (5) investigate whether integration with genetic data could uncover biological mechanisms underlying atherosclerotic disease using genome-wide association study (GWAS) and Mendelian Randomization (MR).

## Methods

### Study population

In this study, we utilized data from the UKB for the primary analysis and the BiDirect Study (Münster, Germany) for external validation. The UKB is a large-scale prospective cohort study that recruited 502,422 individuals aged 40 to 69 at baseline from across the United Kingdom between 2006 and 2010^39^. Participants underwent detailed assessments, which included comprehensive data collection through questionnaires, physical measurements, and biological sample collections.

Following a baseline visit between 2006 and 2010, a total of 82,340 individuals returned for a follow-up imaging visit starting in 2014, which included a carotid ultrasound. A total of 177,757 raw images from 19,768 individuals were released by the UKB and used in this study (**Supplementary Figure 1**). Four anatomic views of the distal common carotid artery and the bifurcation were available for each side for nearly every UKB participant who underwent a carotid ultrasound: images along the main longitudinal axis, images along the short axis, and images along the main longitudinal axis at two different angles for each artery, which were used by the analysts for cIMT quantification. Our study focused on images from 19,507 UKB participants derived along the main longitudinal axis. Eight individuals withdrew from the study post-recruitment (field 190), resulting in a total sample size of 19,499. For participants with repeat imaging visits, only the ultrasound data from the first visit were retained for analysis.

For external validation of the detection model, we used carotid ultrasound images from the BiDirect Study, a twelve-year monocentric prospective cohort study established to explore the relationship between depression and subclinical atherosclerosis^40^. The study recruited patients with clinically diagnosed depression or cardiovascular disease, as well as a population control group. The study participants underwent a standardized carotid ultrasound examination for assessment of cIMT in the context of their baseline and follow-up visits. For the purposes of our study, we utilized carotid ultrasound images along the longitudinal axis from 2,105 patients with available ultrasound data at baseline. The ultrasound examinations in BiDirect were conducted with a different device (Acuson X300, Siemens) than the one used in the UKB (CardioHealth Station, Panasonic) and followed a different protocol.

### Pre-processing

The flowchart for extracting the carotid ultrasound imaging data for analysis from the UKB participants is summarized in **Supplementary Figure 1**. After developing an algorithm (**Supplementary Figure 2**) that automatically detects images along the longitudinal axis, we extracted 45,210 long-axis images without cIMT measurements from the left and right arteries of 19,507 individuals. The obtained images were cropped to a size of 480×448 pixels to retain only the ultrasound image while maintaining the original resolution. After excluding participants who withdrew from the study and retaining only the images from the first ultrasound visit, a total of 19,362 left and 19,370 right carotid images from 19,499 individuals remained for analysis.

In order to enhance contrast and reduce noise in the images, we applied two functions from the *OpenCV* v. 4.7.0 library^41^: median blur filtering (ksize = 5) and Contrast Limited Adaptive Histogram Equalization (clipLimit = 2.0, tileGridSize =(8, 8)), respectively, to facilitate the manual segmentation process. These processing steps were applied to the full sample of images in this study. Plaques were manually annotated by two medical doctors with postgraduate training in vascular imaging and subsequently validated by a doctor certified in carotid ultrasound imaging. *Label Studio* version *1.8.2* (https://github.com/HumanSignal/label-studio) was used to segment plaques on the ultrasound images. The edge coordinates of the segmentation masks were used to obtain the bounding boxes. Plaques were defined, according to standards, as focal protrusions in the arterial lumen with a thickness >50% of the surrounding carotid intima-media thickness^42^. If multiple longitudinal images were available for the same artery, those in which the model detected a plaque were prioritized, or a random image was used if plaques were found in more than one.

For the vast majority of participants in the UKB imaging cohort, only one image along the longitudinal axis of each distal common carotid—including the carotid bulb and bifurcation, but not focused on cIMT measurement (without measurement boxes)—was available for plaque screening. In contrast, the were multiple images across the entire common carotid artery for BiDirect participants. To increase comparability between the two cohorts, we selected, for each BiDirect participant a single image from each artery most likely to depict the region closest to the common carotid artery bifurcation. This selection was enabled by developing a lumen segmentation model based on the U-Net architecture with a pretrained ResNet-34 encoder. For this purpose, we manually segmented lumens in 300 randomly selected images, which were split into training, validation, and testing sets comprising 204, 51, and 45 images, respectively. The model was trained using a combined Dice and binary cross-entropy loss function. After achieving strong segmentation performance in the test set (Dice score: 0.946; IoU: 0.906), we post-processed the predicted segmentation masks to smooth borders and eliminate small artifacts (i.e., objects with an area smaller than 10,000 pixels in images sized 480 × 544), improving the IoU further to 0.914. The process for downstream analysis was implemented with the *scikit-image* (v0.21.0) Python package, with functions from the *morphology* and *measure* modules. The lumen segmentation model was then deployed on the full dataset of 17,864 images. After segmenting all images, the binary masks with the predicted lumen were split in half, and the median height of the left and right halves was measured (representing lumen thickness). The image with the largest difference between the median heights was selected for downstream analysis, as this asymmetry was indicative of closeness to the carotid bifurcation, which consistently appeared on the left side of the image. The lumen segmentation model predictions and post-processed masks are presented in **Supplementary Figure 3**.

### Model Development and Deployment

To train a deep learning model for the detection of atherosclerotic plaques, we manually annotated plaques in 680 randomly selected carotid ultrasound images. A plaque was present in 253 of these images. We performed transfer learning with fine-tuning, which involves selecting a model pre-trained on a large dataset of natural images and then re-training it on a new dataset. This approach allows for adjusting the model’s weights and biases to better suit the task related to the new dataset. Here, we employed the YOLOv8l^43^ model for object detection, pre-trained on over 330,000 images, and re-trained it on our dataset of 680 carotid ultrasound images. The YOLOv8 object detection algorithm generates bounding boxes to indicate the locations of objects of interest. The images with manually annotated plaques were randomly divided into training, validation, and test sets in a 0.725/0.125/0.15 ratio, resulting in 103 images allocated to the test set. This distribution maximized the training set while ensuring a sufficient number of images for assessing model performance in the test set. To enhance model generalizability and predictive power, we randomly selected 50% of the training set (490 images) and applied various augmentation techniques from *Albumentation*s^44^ Python library: GridDistortion (p=0.15), RandomBrightnessContrast ((0,0.5),(0,0.5)), HorizontalFlip(p=0.2), GaussNoise(p=0.15), and RandomSizedCrop (min_max_height=(384, 384), p=0.4). These augmentations increased the variability of the training set, making the model more invariant to noise and other distortions. This augmented dataset, along with the rest of the images, was processed in batches for further augmentation within the YoloV8 framework, as detailed below.

The model was trained with a batch size of 44 images. All tuned and fixed model parameters are listed in **Supplementary Table 1**. The initial learning rate (*lr0*) was set to 0.001, which is ten times lower than the default (0.01), as training was initiated from a pretrained model. A cosine learning rate scheduler was applied to improve convergence. Early stopping with a patience of five epochs was used to prevent overfitting. This means training would stop after five consecutive epochs without improvement. The patience value was chosen based on an initial exploratory run using a preliminary training-validation split, during which we plotted the total training and validation loss curves. In the final run, training concluded after 14 epochs, with peak performance observed at the ninth epoch. The following loss function parameters were selected based on a grid search (**Supplementary Table 1**): distribution focal loss (DFL) = 2.5, box loss = 10 for bounding box regression, and binary cross-entropy loss (CLS) = 1.1. Default augmentation techniques in the YOLOv8 framework were partially suppressed due to prior augmentation efforts; specifically, mosaic, copy-paste, shear, close mosaic, flip up-down, and mix-up were disabled. However, flip left-right (p=0.1), degrees (10), HSV-Saturation (hsv_s: 0.05), HSV-Value (hsv_v: 0.05), translate (0.1), and scale (0.1) were retained. Training was conducted using an NVIDIA QUADRO RTX 5000 GPU (16 GB). *PyTorch*^45^ version *1.12.1* and the *Ultralytics*^43^ framework version *8.1.16* were used for model development.

We evaluated the model’s performance by training it on several subsets of our input development dataset (training + validation) using 5-fold cross-validation. The consistent performance metrics, with minimal variations in precision and recall, indicated no significant signs of overfitting (**Supplementary Figure 4**).

### Model fine-tuning for the BiDirect dataset

To fine-tune our model for the images of the BiDirect cohort that were derived following a different scanning protocol, we manually annotated a set of 600 images using the same plaque definition as for the UKB sample. Of these images, 29.8% (179) contained plaques. The dataset was split into training (336 images), validation (144 images), and test (120 images), sets.

Fine-tuning was implemented using the previously developed model on the UK Biobank images, with a reduced learning rate of 0.0001, ‘warmup_epochs’ set to 0, and the ‘pretrained’ option enabled. No additional augmentation was employed beyond the one natively implemented in the *Ultralytics* framework, with the same parameters used in training on the UKB images (**Supplementary Table 1**). Loss coefficients were set to the default values (cls = 0.5; dfl = 1.5; box = 7.5). Patience was set to 5 epochs to avoid overfitting. Batch size was set to 44 to fit constraints, and the image size (*imgsz* parameter) was set to 480, as it was for the UKB images.

Model’s classification metrics for detecting plaque-positive images were estimated in a confusion matrix by comparing ground truth annotations with bounding box predictions. A true positive was recorded when both the annotation and the prediction contained a bounding box.

### Demographic and Clinical Variables

In the UKB, age at carotid ultrasound assessment was derived by subtracting the participant’s date of birth (field 33) from the carotid ultrasound assessment visit date (extracted from the manifest files linked to the ultrasound images). Ethnicity for the PCE risk score calculation was determined by field 21000. Observations with missing data or responses of “Do not know” or “Prefer not to answer” were encoded as “Other”. Pre-existing CVD was defined based on self-reported history (UKB field 20002, with 1075 - heart attack/myocardial infarction, 1081 - stroke, 1583 - ischemic stroke, 1491 - brain haemorrhage); general practice records (131298, 131300, 131302, 131368, 131366); hospital data records (defined using ICD-10 codes I20-I25, I60-I61, and ICD-9 codes 410-412, 429-431, 434, 436, as well as operation codes K40-46, K49, K471, K49, K50, K75, L294, and L295) observed before the initial ultrasound visit. SBP was quantified using fields 4080 and 93 by averaging the observations from each field, followed by taking the mean of the resulting values. Smoking status was categorized as "current" or "other", with missing data (<0.5%) treated as "other" (field 20116). Total cholesterol, LDL and HDL cholesterol levels were obtained from fields 30690, 30780 and 30760, respectively. When available, values from the assessment closest to the ultrasound visit were extracted; otherwise, baseline measurements were used. Missing values were imputed using the multivariate imputation by chained equations method^46^, affecting 12% of HDL-cholesterol values and 6% of total and LDL-cholesterol values, based on other CVD risk factors. Five imputations were conducted, and prediction results were pooled using Rubin’s rules. Diabetes was defined by self-reported cases (UKB field 20002 – codes: 1220, 1222, 1223), use of glucose-lowering medications (field 20003), and hospital records prior to the first carotid ultrasound exam (ICD-9 codes: 250* and ICD-10 codes E10, E11). Information on the use of antihypertensive drugs, diabetes medications, and statins was obtained from field 20003 (**Supplementary Table 2).**

### Assessment of major adverse cardiovascular events

MACE endpoints were defined as follows: myocardial infarction (ICD-10 codes I21, I22 from hospital inpatient data, UKB data-fields 131298 and 131300), stroke (ICD-10 codes I60–64 from hospital inpatient data; UKB data-fields 131368 and 131366), or death due to any cardiovascular cause (defined as the cause of death with an ICD-10 code starting with ’I’, extracted from the death registry). The date of the first episode observed after the carotid ultrasound assessment was considered as the date of the event of interest.

### Carotid intima-media thickness

We compared three cIMT measurements between UKB participants with and without carotid plaque, as predicted by the model. Measurements of mean and maximum cIMT were obtained using UKB data fields 22670–22681, as described by Strawbridge et al^47^.

- cIMT mean: average of the mean values from four mean cIMT measurements (two angles for each carotid artery: left and right).
- cIMT max: The maximum value of the cIMT measurements across both arteries.
- cIMT mean-max: The mean of the maximum cIMT values per artery.

All the obtained values were log-transformed for the analysis. For the analysis comparing the two arteries, individuals with missing cIMT measurements for either the left or right artery were excluded, resulting in a total of 18,497 individuals included in this analysis. The analysis was also repeated separately for the right and left arteries. For the right artery, data were available for 18,803 individuals with no missing measurements; for the left artery, data were available for 18,703 individuals with no missing measurements. Statistical tests were performed on the specified number of individuals in each subset.

For the BiDirect participants, mean cIMT measurements were available for 1,974 and 1,956 individuals for the left and right arteries, respectively, out of 2,105 participants. Missing values were excluded from the comparative analysis.

### Logistic regression for plaque presence and Cox regression for future events

To estimate the associations between model-derived plaque presence and CVD risk factors, we applied logistic regression, using plaque presence as the outcome variable. A history of hypertension was defined by the use of antihypertensive treatment at the time of the carotid ultrasound assessment. Definitions of other clinical and demographic variables are described above.

For each subset of the cohort (full sample, primary events with and without individuals on statin therapy), two separate Cox regression models were constructed: one with plaque presence as a binary variable and another with the count of plaques as a categorical variable with three levels (no plaques as the reference, one plaque, and two or more plaques). The time variable in the survival analysis was calculated as the duration from the first ultrasound visit to the event of interest or to the censoring date, which included date of death from causes other than CVD or the date at the last observed event (2022-10-30) available at the time of the analysis. Controls were censored at the time point of the latest observed event. The fitted Cox regression models included all vascular risk factors included in the PCE (age, sex, SBP, smoking status, history of diabetes, antihypertensive therapy, total cholesterol, and HDL cholesterol), as well as statin usage. Interaction with sex was evaluated by including an interaction term in the Cox models. Ancestry was not included due to its very low variance in our sample (**Table 1**). The proportional hazards assumptions were tested with the scaled Schoenfeld residuals, and no violation of the assumptions was detected.

**Table 1.**
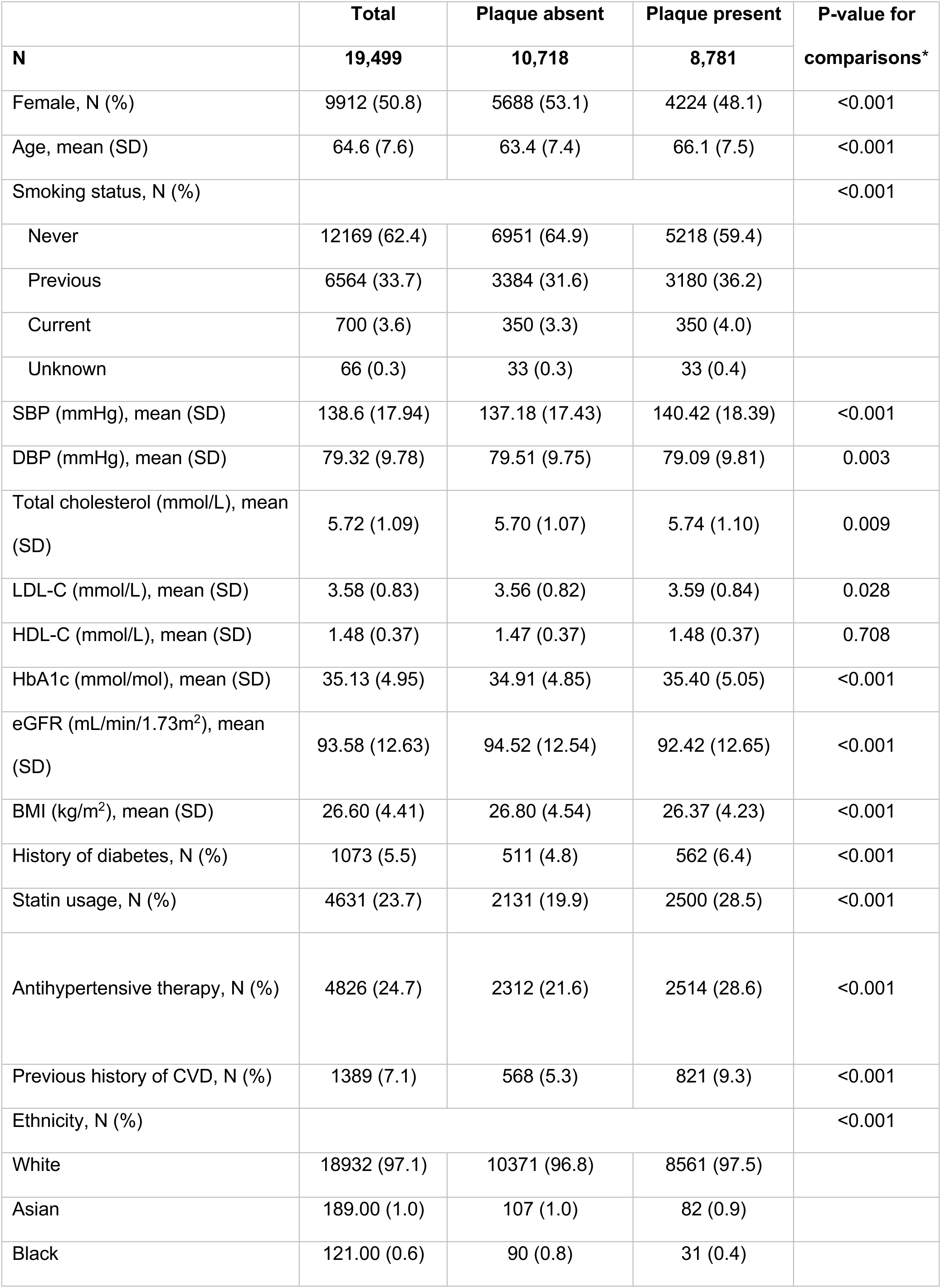
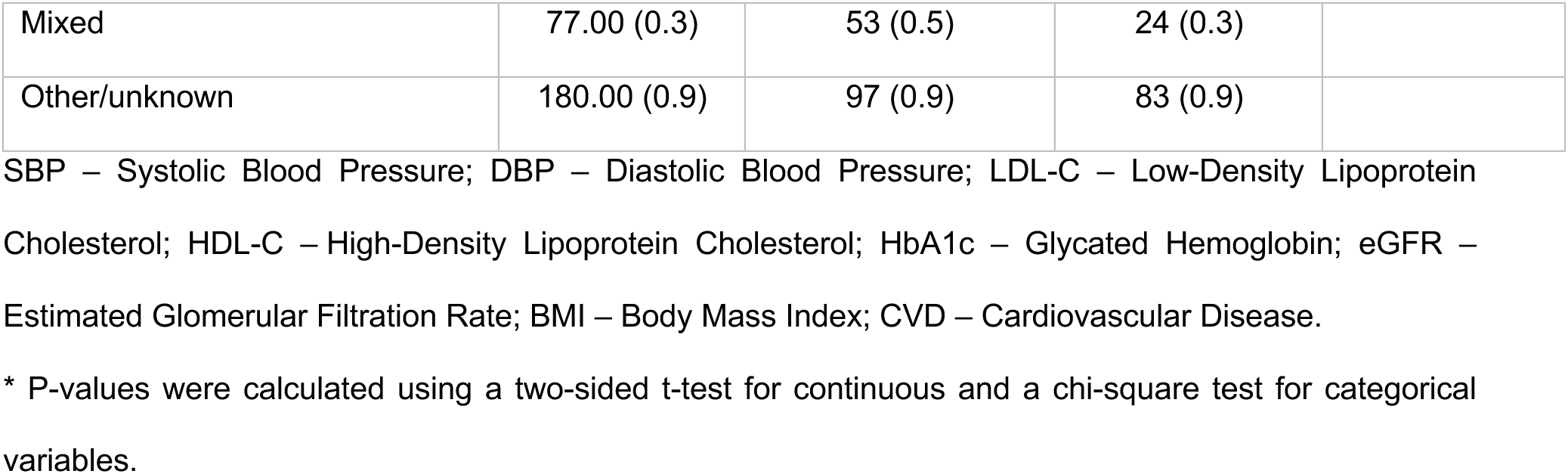
Characteristics of the study cohort at the time of initial carotid ultrasound assessment.

|  | Total | Plaque absent | Plaque present | P-value for |
| --- | --- | --- | --- | --- |
| N | 19,499 | 10,718 | 8,781 | comparisons* |
| Female, N (%) | 9912 (50.8) | 5688 (53.1) | 4224 (48.1) | <0.001 |
| Age, mean (SD) | 64.6 (7.6) | 63.4 (7.4) | 66.1 (7.5) | <0.001 |
| Smoking status, N (%) |  |  |  | <0.001 |
| Never | 12169 (62.4) | 6951 (64.9) | 5218 (59.4) |  |
| Previous | 6564 (33.7) | 3384 (31.6) | 3180 (36.2) |  |
| Current | 700 (3.6) | 350 (3.3) | 350 (4.0) |  |
| Unknown | 66 (0.3) | 33 (0.3) | 33 (0.4) |  |
| SBP (mmHg), mean (SD) | 138.6 (17.94) | 137.18 (17.43) | 140.42 (18.39) | <0.001 |
| DBP (mmHg), mean (SD) | 79.32 (9.78) | 79.51 (9.75) | 79.09 (9.81) | 0.003 |
| Total cholesterol (mmol/L), mean (SD) | 5.72 (1.09) | 5.70 (1.07) | 5.74 (1.10) | 0.009 |
| LDL-C (mmol/L), mean (SD) | 3.58 (0.83) | 3.56 (0.82) | 3.59 (0.84) | 0.028 |
| HDL-C (mmol/L), mean (SD) | 1.48 (0.37) | 1.47 (0.37) | 1.48 (0.37) | 0.708 |
| HbA1c (mmol/mol), mean (SD) | 35.13 (4.95) | 34.91 (4.85) | 35.40 (5.05) | <0.001 |
| eGFR (mL/min/1.73m<sup>2</sup>), mean (SD) | 93.58 (12.63) | 94.52 (12.54) | 92.42 (12.65) | <0.001 |
| BMI (kg/m<sup>2</sup>), mean (SD) | 26.60 (4.41) | 26.80 (4.54) | 26.37 (4.23) | <0.001 |
| History of diabetes, N (%) | 1073 (5.5) | 511 (4.8) | 562 (6.4) | <0.001 |
| Statin usage, N (%) | 4631 (23.7) | 2131 (19.9) | 2500 (28.5) | <0.001 |
| Antihypertensive therapy, N (%) | 4826 (24.7) | 2312 (21.6) | 2514 (28.6) | <0.001 |
| Previous history of CVD, N (%) | 1389 (7.1) | 568 (5.3) | 821 (9.3) | <0.001 |
| Ethnicity, N (%) |  |  |  | <0.001 |
| White | 18932 (97.1) | 10371 (96.8) | 8561 (97.5) |  |
| Asian | 189.00 (1.0) | 107 (1.0) | 82 (0.9) |  |
| Black | 121.00 (0.6) | 90 (0.8) | 31 (0.4) |  |
| Mixed | 77.00 (0.3) | 53 (0.5) | 24 (0.3) |  |
| Other/unknown | 180.00 (0.9) | 97 (0.9) | 83 (0.9) |  |

- 1 SBP – Systolic Blood Pressure; DBP – Diastolic Blood Pressure; LDL-C – Low-Density Lipoprotein - 2 Cholesterol; HDL-C – High-Density Lipoprotein Cholesterol; HbA1c – Glycated Hemoglobin; eGFR – - 3 Estimated Glomerular Filtration Rate; BMI – Body Mass Index; CVD – Cardiovascular Disease. \* P-values were calculated using a two-sided t-test for continuous and a chi-square test for categorical variables.

All analyses were conducted using R software, version 4.4.0. We considered two-sided p-values (unless otherwise specified) less than 0.05 to be statistically significant. The category-free NRI was calculated using the *nricens* v.1.6^48^ package, with confidence intervals and p-values based on 1,000-fold bootstrap standard errors. The IDI metric was calculated using the *survIDINRI* library v.1.1-2^49^. Harrell’s C-statistic, along with its 95% confidence interval, was used to evaluate the discriminative ability of the time-to-event models^50^. Comparisons between models based on the C-statistic were conducted using the *CsChange* package^51^.

### PCE risk estimation

The calculation of the PCE risk score was performed using published equations^6^. PCE eligibility included the following criteria:

1) 40≤Age≤79,
2) 130 ≤ Total cholesterol ≤320 mg/dL;
3) 20 ≤ HDL-cholesterol ≤ 100 mg/dL;
4) 90 ≤ Systolic blood pressure ≤ 200 mmHg

To enable fair model comparison and justify the recommended threshold usage, the original PCE was calibrated to the UKB population. This recalibration was achieved by fitting calculated log-hazards from the published PCE coefficients in a Cox regression stratified by sex to obtain recalibrated probabilities. The calibration of the predicted risk values from the original and recalibrated models was assessed using Greenwood–Nam–D’Agostino statistics and the integrated calibration index^52,53^. Calibration plots for the PCE are presented in **Supplementary Fig. 5**.

The plaque variables were incorporated into the PCE, with plaque presence as a binary variable and plaque count as an ordinal variable, as previously described^54^. Briefly, the recalculated risk is derived from the relative risk estimate for the novel risk factor, the baseline risk, and the prevalence of the novel risk factor. As an example, the presence of plaque was incorporated using Equation (1), adapted from Kooter et al^54^:

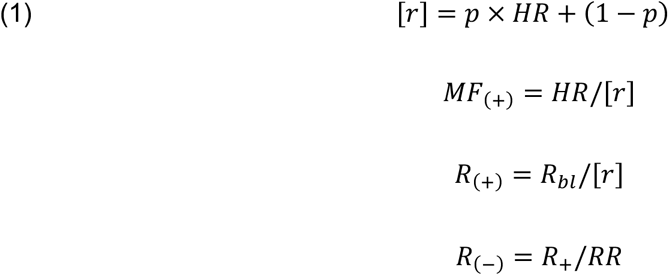

where [r] represents the weighted mean risk; p is the plaque prevalence; HR is the hazard ratio for plaque presence; MF_(+)_ is the multiplication factor; R_(+)_ is recalculated risk in the presence of plaque; R_bl_ is the baseline risk estimated from the baseline hazard and log-HRs estimated from the fitted Cox model ; and R_(−)_ is the recalculated risk in the absence of plaque.

HRs for both plaque presence and plaque count, used to enhance the recalibrated PCE risk model with plaque information, were estimated from a Cox regression model fitted on the sub-cohort eligible for PCE risk assessment. The model was adjusted for sex, age, HDL and total cholesterol levels, antihypertensive drug use, smoking status, SBP, and statin use at the time of the initial ultrasound assessment. Plaque information was incorporated separately for males and females, with prevalence estimates calculated for each group.

Model calibration was assessed using the *survival.calib*^55^ package in R. Reclassification tables and NRI statistics were calculated using the *PredictABEL* v.1.2-4^56^ library in R.

### Genome-wide association study and meta-analysis

Genomic quality control in the UKB was performed as previously described^57^. For the GWAS analysis, *Regenie*^58^ v3.3 was employed. In step 1, we used directly genotyped variants with MAF > 1%, <10% missingness, Hardy–Weinberg equilibrium test *P* > 1 × 10^−1^^5^, and a minor allele count > 100. Age at the time of the ultrasound exam, along with sex, the first 10 genetic principal components, and the genotyping chip were used as covariates. Fixed-effect meta-analysis was conducted with *METAL*^59^, using effect size estimates and standard errors (option *SCHEME STDERR*). Between-study heterogeneity was assessed using Cochran’s Q statistic as implemented in *METAL*. Significant heterogeneity was defined as a Cochran’s Q-derived p-value of less than 0.05. Results were clumped using the *clump_data* function of *TwoSampleMR*^60,61^ R package version 0.5.6) at an *r*^2^<0.001 based on the European 1000 Genomes Project reference panel with 10,000 kb window^62^. Results were then visualized in a Manhattan plot, which was constructed using the *gwaslab* Python package^63^.

### Mendelian randomization

MR aims to estimate the causal effect of an exposure on an outcome using genetic variants as instrumental variables, under three assumptions: (1) the instrument is robustly associated with the exposure; (2) the instrument is independent of confounders of the exposure–outcome relationship; and (3) the instrument affects the outcome only through the exposure (no horizontal pleiotropy). Violations of these assumptions introduce bias. Summary-level data sources for exposures, along with their descriptions, are provided in **Supplementary Table 3**. Two-sample MR was conducted using the *TwoSampleMR v0.6.7* package in R. The instrumental variable for each exposure was constructed by selecting SNVs from summary statistics files with a significant association (p < 5×10⁸), followed by clumping for linkage disequilibrium at an r² < 0.001 threshold within a 10,000 kb window. R² and F-statistics were calculated for each SNP to assess instrument strength and evaluate potential weak instrument bias (**Supplementary Table 4**). Only SNPs with F-statistics > 10 were retained for subsequent analytical procedures to ensure adequate instrument strength. For IL-6 receptor-mediated signaling activity, the genetic instrument was constructed as previously described^64^, using beta estimates derived from a cohort that excluded the UKB to minimize bias. Main association estimates were derived using random-effects IVW analysis. To account for potential bias due to horizontal pleiotropy, sensitivity analyses were performed using both MR-Egger and the weighted median estimator, as these methods are known to be more robust against pleiotropic effects^65,66^. Heterogeneity and horizontal pleiotropy of the genetic instruments were estimated using the *mr_heterogeneity* and *mr_pleiotropy_test* functions from the *TwoSampleMR* package. When MR-Egger indicated significant pleiotropy, we utilized the Mendelian Randomization Pleiotropy RESidual Sum and Outlier (MR-PRESSO) method from the *MRPRESSO*^67^ *v1.0* package in R to identify and exclude significant pleiotropic instrumental variables (P < 0.05), which were considered as “outliers”. Subsequently, IVW, weighted median, and MR-Egger analyses were carried out on the outlier-corrected models. Continuous traits (HbA1c, BMI, waist-to-hip ratio, LDL-cholesterol, HDL-cholesterol) were rank-inverse-normalized in source GWASs; inverse-variance-weighted (IVW) estimates represent effects per SD increment. Blood pressure coefficients were scaled by a factor of 10, expressing effects per 10 mmHg. Binary exposures (smoking initiation, type 2 diabetes) modeled on the log-odds scale were multiplied by 0.693, representing two-fold odds differences. IL-6 signaling was assessed using a 26-variant IL6R instrument per unit increase in natural logarithm-transformed C-reactive protein. False discovery rate (FDR) correction was applied to the IVW estimates to control for multiple testing.

We conducted the MR study in accordance with the guidelines of Strengthening the Reporting of Observational Studies in Epidemiology using Mendelian Randomization (STROBE-MR).^68,69^ The STROBE-MR checklist is provided in **Supplementary Table 5.**

### Data Availability Statement

The UKB provides an accessible research resource, available to researchers upon submitting a research proposal at https://www.ukbiobank.ac.uk. The GWAS data on UKB and the GWAS meta-analysis results obtained in this study are available in the GWAS Catalog (https://ftp.ebi.ac.uk/pub/databases/gwas/summary_statistics/GCST90454001-GCST90455000/) under accession numbers GCST90454354 and GCST90454353, respectively. Additionally, the carotid plaque phenotypes derived from the developed model will be returned to UKB for use in future studies. The list of GWAS used for MR analyses are presented in **Supplementary Table 3**. Individual-level data of the BiDirect cohort can be obtained by researchers from KB (https://medizin.uni-muenster.de/en/epi) following a written transfer agreement.

The code used for this study will be available on GitHub upon publication of this manuscript.

### Ethical Considerations

The UK Biobank has obtained approval from the Northwest Multi-Center Research Ethics Committee. All participants have provided written informed consent. The Bidirect study received ethical approval from the Ethics Committee of the University of Münster and the Westphalian Chamber of Physicians in Münster, Germany (# 2009-391-f-S). Written informed consent was obtained from all participants.

## Results

### Study population

A total of 177,757 images from 19,499 participants who underwent carotid ultrasound during the first imaging visit of the UKB were available for analysis (**Supplementary Fig. 1**). The protocol for carotid artery examination has been described previously^70^. For the current study, we used 38,732 images obtained in the longitudinal axis of the left and right distal common carotid artery and the bifurcation, allowing for the assessment of plaque presence along the vessel wall. The demographic and medical characteristics of the study participants are presented in **Table 1**. The mean age at the time of the carotid ultrasound examination was 64.6 years (SD = 7.59), and 50.8% of the participants were female. A total of 1,381 (7.1%) study participants had a baseline diagnosis of atherosclerotic CVD. A comparison between UKB participants with carotid ultrasound and the rest of the UKB cohort revealed a lower prevalence of CVD risk factors (**Supplementary Table 6**).

### Plaque detection model

To train a deep learning model for the detection of atherosclerotic plaques, we manually annotated plaques in 680 randomly selected carotid ultrasound images. Plaques were defined as focal protrusions in the arterial lumen with a thickness greater than 50% of the surrounding carotid intima-media thickness^42^. A plaque was present in 253 of these images. We performed transfer learning with fine-tuning by employing a pre-trained YOLOv8^43^ object detector as the foundation for developing the plaque detection model. The YOLOv8 object detection algorithm generates bounding boxes to indicate the locations of objects of interest. The images with manually annotated plaques were randomly divided into training, validation, and test sets in a 0.725/0.125/0.15 ratio. This distribution maximized the training set while ensuring a sufficient number of images for assessing model performance in the test set. We evaluated the model’s performance by training it on several subsets of our input development dataset (training + validation) in 5-fold cross-validation. The consistent performance metrics, with minimal variations in precision and recall, indicated no significant signs of overfitting (**Supplementary Fig. 4**).

After training, the performance of the model was evaluated in a blind test set of 103 images, of which 38 contained at least one plaque (53 plaques in total). The model achieved high classification metrics for plaque presence at the image level, with an accuracy, sensitivity, specificity, and Positive Predictive Value (PPV) of 89.3%, 89.5%, 89.2%, and 82.9% (**Figure 1A)**, respectively, at an iteratively tuned confidence score threshold of 13%. The confidence score measures the model’s certainty that a box contains an object of interest and was tuned to optimize and balance accuracy, sensitivity, and specificity. The Mean Average Precision at an Intersection over Union (IoU) threshold of 50% (mAP@50) was 68.4%, indicating the precision with which the model can localize objects with at least 50% overlap with the ground truth. The model’s detection precision and recall were 70.4% and 71.7%, respectively. Prediction examples are illustrated in **Figure 1B** and **Supplementary Fig. 6**. Overall, the model correctly identified 42 of 53 plaques (79.2%) in 38 plaque-positive images in the test set (**Supplementary Fig. 7**).

**Figure 1.**
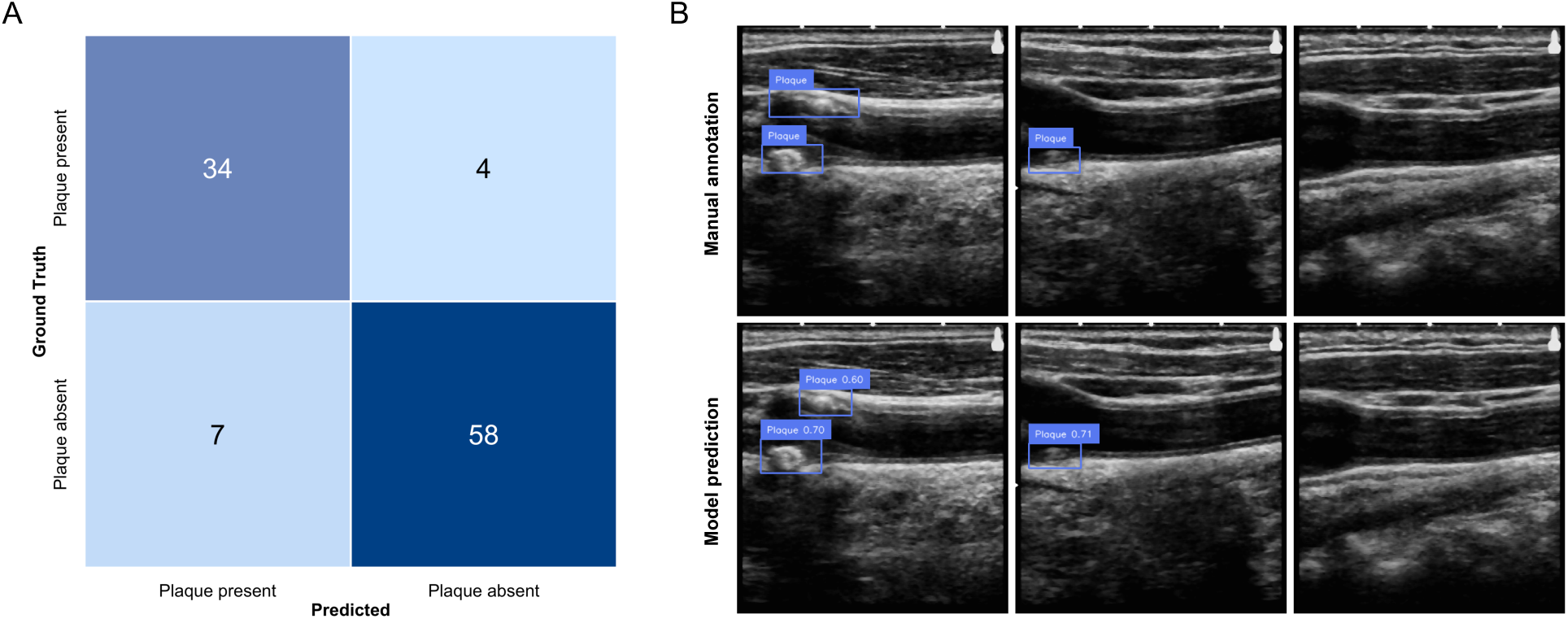
Development and performance of the plaque detection model. **A**. Confusion matrix for model’s classification performance for plaque presence at each image. The confusion matrix is based on comparing the presence of plaque in the annotations with the model’s predictions. Specifically, if there is a plaque annotation for an image and the prediction contains a bounding box, then the prediction is annotated as true positive. **B.** Examples of model predictions as compared to manual annotations for three UK Biobank participants in the test set. Each image depicts the longitudinal view of the common carotid artery extending toward the bifurcation area (the left part of the images).

To assess extent to which the augmentation parameters affect the results, we conducted a post hoc sensitivity analysis using varying augmentation probabilities for *GridDistortion* (0.05, 0.15, 0.25), *HorizontalFlip* (0.1, 0.2, 0.3), and *RandomSizedCrop* (0.3, 0.4, 0.5). The detection metrics ranged from 0.52 to 0.85 for precision, 0.45 to 0.66 for recall, and 0.50 to 0.74 for mAP@50 on the test set with a default confidence threshold of 50%. The results indicate that image augmentation may affect the outcomes, but the developed model achieved optimal performance under the selected augmentation scheme.

### Prevalence and risk factors of carotid plaques in the UK Biobank

Next, we deployed the model on all available long-axis carotid ultrasound images from the UKB cohort (38,732 images, 19,499 individuals, **Supplementary Fig. 1**). This deployment allowed us to extract data on plaque phenotypes, including plaque presence and the count of plaques in either artery for each individual. The count of plaques was determined by the number of predicted bounding boxes in each image.

Overall, the model detected at least one plaque in 45% of UKB participants who underwent a carotid ultrasound examination. In 14% of participants, the model detected at least two plaques, and in 3.1%, three or more plaques across both arteries. The prevalence of plaques in the left and right carotid arteries is presented in **Figure 2A**. As a quality control step for the model predictions, we explored whether plaque presence was associated with cIMT, which was quantified and documented for each individual at the time of the imaging assessment. Indeed, cIMT was consistently higher for individuals for whom our model predicted a plaque (Wilcoxon test p < 10^-60^ for maximum, mean, and mean of maximum cIMT measurements). Similar results were obtained for the left and right arteries separately (**Supplementary Fig. 8)**.

**Figure 2.**
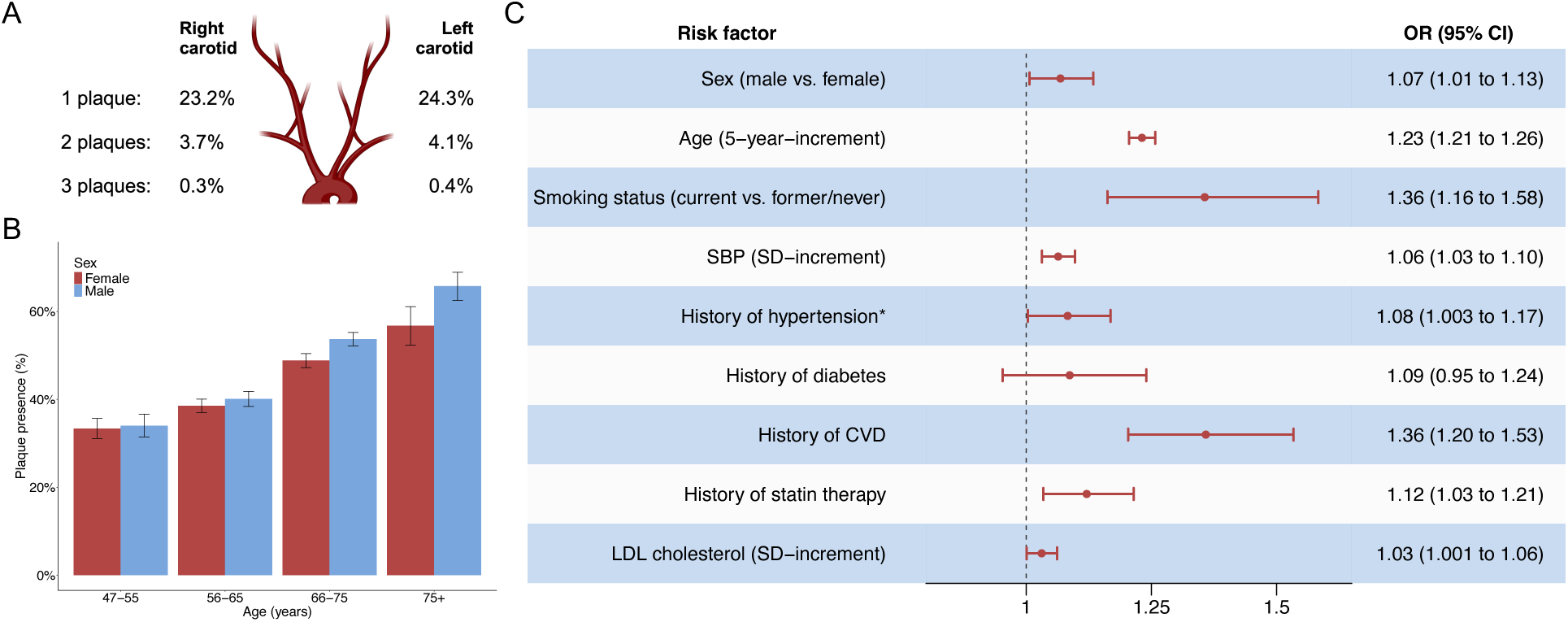
Prevalence and predictors of carotid plaque in the UK Biobank. **A**. Distribution of plaques in the left and right carotid arteries. **B**. Percentage of plaque presence across age and sex groups. Error bars represent 95% confidence intervals. **С.** Forest plot of the associations of demographic and vascular risk factors with the odds of carotid plaque presence, as derived by a multivariable logistic regression model that includes all variables in the figure. The results are presented as odds ratios (OR) and 95% confidence intervals (CI). SBP – systolic blood pressure; CVD – cardiovascular disease; LDL – low-density lipoprotein cholesterol; SD – standard deviation. * History of hypertension is defined by the use of antihypertensive medication

Plaques were more common in male participants (47.5% vs. 42.6%; two-proportions z-test, p = 5.7 x 10^-12^) and plaque prevalence was significantly associated with older age, increasing from 31.1% in participants aged 45–54 years to 62% in participants aged 75 years or older (Cochran-Armitage trend test, p = 1.12 x 10^-88^, **Figure 2B**). In a multivariable logistic regression model, male sex, older age, current smoking, higher systolic blood pressure (SBP), history of hypertension, pre-existing CVD, use of statins, and higher Low-Density Lipoprotein (LDL) cholesterol levels were all significantly associated with plaque presence (**Figure 2C**).

### External validation of plaque detection model in the BiDirect study

To ensure external validity of our plaque detection model, we conducted validation in the BiDirect cohort. Baseline characteristics of the BiDirect participants included in this study are presented in **Supplementary Table 7.** After fine-tuning the developed model on the BiDirect images, we evaluated it on a test set of 120 images, which included 40 plaque-positive images. At a confidence threshold of 0.13, the fine-tuned model reached high image-level classification metrics for plaque presence (accuracy 85.8%, sensitivity 77.5%, specificity 90%, and PPV 79.5%, **Figure 3A**). The mAP@50 of the fine-tuned model was 65.1%. The model detected 29 of 45 plaques in 40 plaque-positive images (**Supplementary Figure 9**). Examples of the model’s predictions are shown in **Figure 3B**. In a small subset of examinations (n = 64) annotated by a sonographer assessing full image loops, our model, based on a single image, showed high agreement in correctly detecting 90% of right and 86% of left carotid plaques.

**Figure 3.**
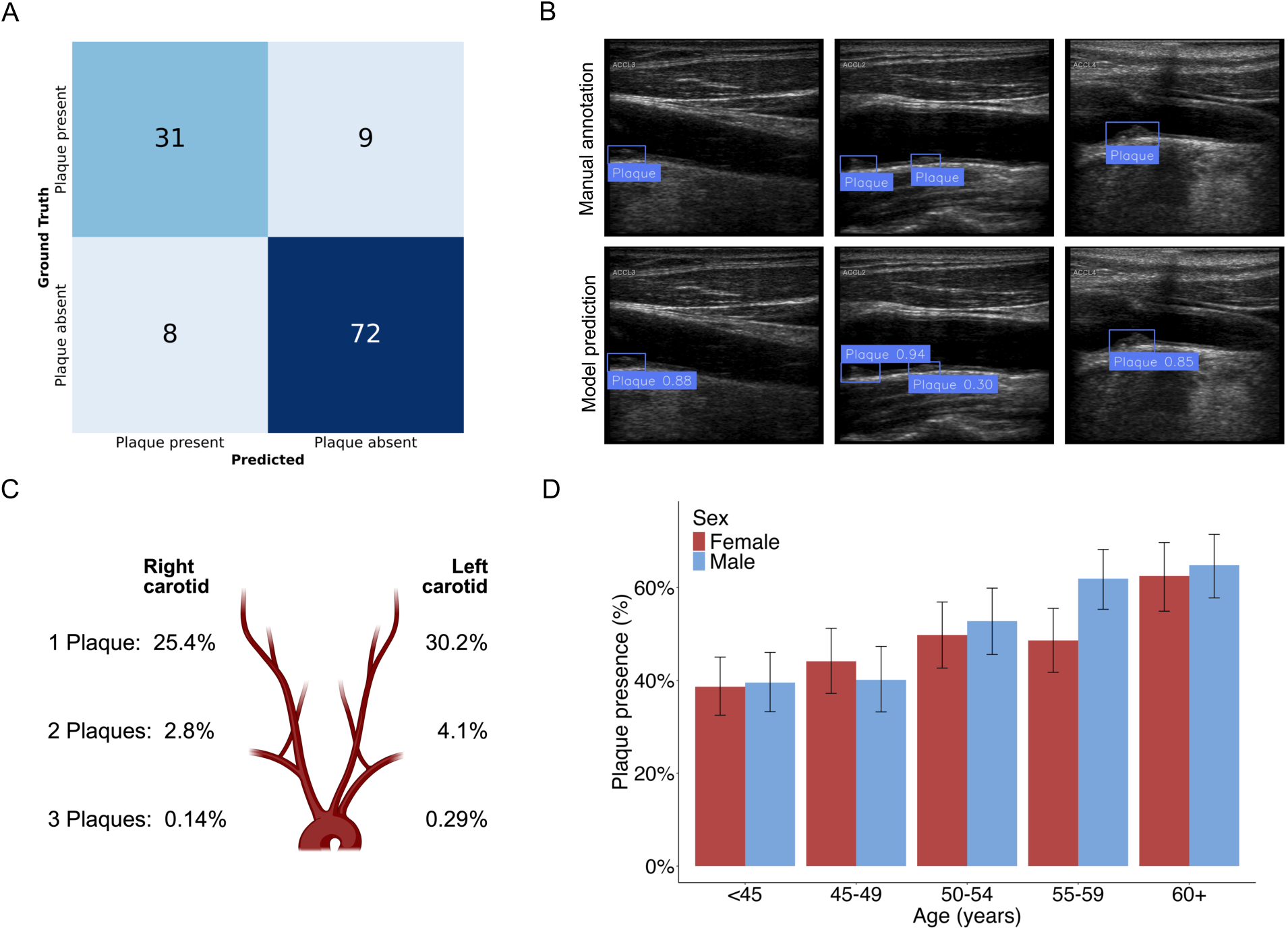
Performance of the fine-tuned plaque detection model in the BiDirect sample. **A**. Confusion matrix for model’s classification performance for plaque presence at each image. **B.** Examples of model predictions as compared to manual annotations for four BiDirect participants in the test set. **C.** Distribution of plaques in the left and right carotid arteries. **D.** Percentage of carotid plaque across age and sex groups. Error bars represent 95% confidence intervals.

In the full dataset of 2,105 patients, the model detected at least one plaque in 1,049 patients (49.8%). In 12.9% of study participants, plaques were detected bilaterally. The prevalence of plaques was 34.6% in the left artery and 28.3% in the right artery (**Figure 3C**). Plaque presence was significantly associated with older age, increasing from 39% in participants younger than 45 years to 63.7% in those aged 69 years or older (Cochran–Armitage trend test, p = 5.31 × 10⁻¹⁶, **Figure 3D**). Plaque prevalence was higher in the CVD group (59.0%) than in the depression (45.7%) and population control groups (51.0%; one-sided two-proportion z-tests with Holm adjustment: p = 6.3×10⁻⁵ and p = 0.015, respectively). BiDirect participants with predicted plaques had higher mean cIMT values in both the left and right arteries (Wilcoxon p-values: 9.2 × 10⁻¹⁵ for the left and 4.6 × 10⁻⁸ for the right carotid artery; **Supplementary Figure 10**).

### Associations of plaque phenotypes with the risk of future cardiovascular events

To estimate the effects of the presence and count of plaques in carotid ultrasound on the risk of future major adverse cardiovascular events (MACE), we conducted a survival analysis. Following the first imaging visit, UKB participants with available ultrasound images were followed up for a median of 55 months (range 1-80 months). During this time interval, a total of 430 individuals experienced a MACE, defined as myocardial infarction, stroke or death due to any cardiovascular cause. Of these, 335 were first-ever events among 18,110 participants (1.8%) without a history of CVD at the time of the ultrasound examination, while 95 were secondary events among 1,389 participants (6.8%) with an existing history of CVD.

Kaplan-Meier estimates indicated a higher incidence rate of MACE among individuals with carotid plaques compared to those without plaques, demonstrating a dose-response pattern of higher incidence with an increasing plaque count (log-rank test p-value for all pairwise comparisons < 0.05; **Figure 4**). After adjusting for conventional cardiovascular risk factors in Cox regression models, plaque presence was significantly associated with future risk of MACE (Hazard Ratio (HR) 1.42, 95% CI: 1.16–1.73). The plaque count per individual showed a dose-dependent association with future CVD risk (HR for 1 plaque vs. no plaque = 1.30, 95% CI: 1.04–1.63; HR for 2 or more plaques vs. no plaques = 1.62, 95% CI: 1.27–2.07). These associations were consistent in the subgroup of individuals without an existing history of CVD, as well as in those who had neither a history of CVD nor statin use (**Supplementary Figure 11**). There was no evidence of an interaction with sex (p = 0.139). The HRs were comparable when analyzing individual MACE components, albeit with wider 95% CIs–likely due to lower statistical power (195 myocardial infarction and 172 stroke cases; **Supplementary Figure 11**).

**Figure 4.**
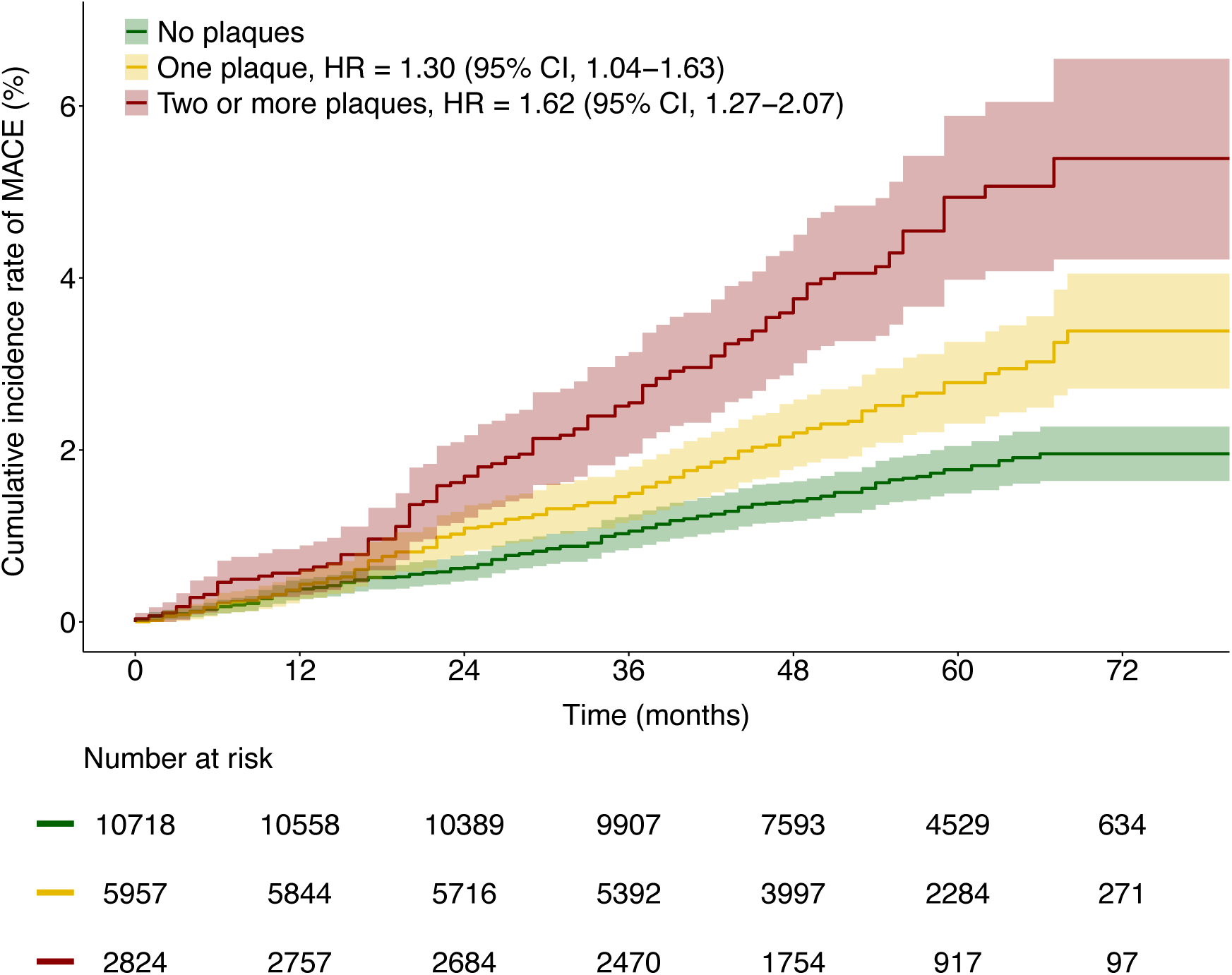
Survival curves of cumulative major adverse cardiovascular event (MACE) rates by total count of carotid plaques predicted by the model. The presented hazard ratios (HRs) were estimated using Cox regression, adjusted for: sex, age, systolic blood pressure, use of statins, history of antihypertensive therapy, current smoking, history of diabetes, HDL cholesterol and total cholesterol levels.

Sensitivity analysis with adjustment for competing risks was conducted using Fine-Gray regression, where deaths from causes other than CVD (203 individuals) were treated as competing risks. Subdistribution hazard ratios (HRs) from the Fine-Gray models were similar to those derived from Cox regression models, showing HRs for both plaque presence (subdistribution HR = 1.42, 95% CI: 1.16–1.73) and plaque count (subdistribution HR for 1 plaque vs. no plaque = 1.31, 95% CI: 1.04–1.64; HR for 2 or more plaques vs. no plaques = 1.61, 95% CI: 1.26–2.05).

### Predictive power of plaque phenotypes

To examine whether assessing carotid plaque phenotypes could improve cardiovascular risk prediction, we compared the fitn, reclassification, and discrimination of prediction models that included conventional vascular risk factors with those that also considered plaque presence and count. Both plaque presence and count significantly improved the overall goodness of fit of a Cox regression model (p < 0.05), as assessed by the log-likelihood ratio test^71^. We observed a reclassification improvement for plaque presence (category-free net reclassification improvement (cfNRI): 0.331, 95% CI, 0.217–0.445) and plaque count (cfNRI: 0.369, 95% CI, 0.260–0.478, **Table 2**). Briefly, NRI quantifies model improvement by the difference between the net proportion of cases for which the new model correctly increases predicted risks and net proportion of controls for which the new model correctly decreases predicted risks^72,73^.

**Table 2.** Metrics of discrimination, reclassification and overall model fit between Cox regression models with and without plaque information.

| Model | Control /<br>Cases | Log-likelihood<br>ratio test p-value | cfNRI<br>[95% CI]<br>(p-value) | IDI<br>[95 % CI]<br>(p-value) | C-index<br>[95% CI] | C-index<br>difference<br>(p-value) |
| --- | --- | --- | --- | --- | --- | --- |
| CVD factors | 19069 /<br>430 | ref | ref | ref | 0.745 [0.723; 0.767] | ref |
| CVD factors<br>+ Plaque<br>presence |  | 4.8x10<sup>-4</sup> | 0.331 [0.217; 0.445]<br>(1.26x10<sup>-8</sup>) | 0.0022 [0.0002; 0.0050]<br>(0.019) | 0.747 [0.725; 0.769] | 0.002 (0.222) |
| CVD factors<br>+ Plaque<br>count |  | 5.6x10<sup>-4</sup> | 0.369 [0.260; 0.478]<br>(2.97x10<sup>-11</sup>) | 0.0024 [2x10<sup>-5</sup>; 0.0063]<br>(0.048) | 0.748 [0.726; 0.770] | 0.003 (0.173) |
| CVD factors | 17775 /<br>335 | ref | ref | ref | 0.744 [0.719; 0.770] | ref |
| CVD factors<br>+ Plaque<br>presence |  | 0.003 | 0.329 [0.206; 0.452]<br>(1.65x10<sup>-7</sup>) | 0.0019 [0.0003; 0.0052]<br>(0.0079) | 0.746 [0.721; 0.771] | 0.002 (0.357) |
| CVD factors<br>+ Plaque<br>count |  | 0.006 | 0.330 [0.209; 0.450]<br>(7.73x10<sup>-8</sup>) | 0.0022 [0.0004; 0.0065]<br>(0.0139) | 0.746 [0.721; 0.772] | 0.002 (0.349) |
| CVD factors | 14283 /<br>234 | ref | ref | ref | 0.761 [0.732; 0.790] | ref |
| CVD factors<br>+ Plaque<br>presence |  | 0.014 | 0.308 [0.159; 0.457]<br>(4.88x10<sup>-5</sup>) | 0.0018 [0.0002; 0.0059]<br>(0.014) | 0.763 [0.734; 0.792] | 0.002 (0.453) |
| CVD factors<br>+ Plaque<br>count |  | 0.039 | 0.308 [0.159; 0.458]<br>(5.31x10<sup>-5</sup>) | 0.0021 [0.0003; 0.0075]<br>(0.0119) | 0.763 [0.734; 0.792] | 0.002 (0.471) |

CVD – cardiovascular disease; cfNRI – category-free net reclassification improvement; C-index – concordance index; IDI – integrated discrimination improvement; ref – reference.

Sensitivity analyses confirmed these improvements in individuals without a history of CVD and statin use (**Table 2**). Despite the strong associations with the future risk of MACE, none of the plaque phenotypes added to conventional risk factors significantly improved model discrimination, as measured by the C-index. Specifically, adding plaque presence to conventional risk factors only slightly changed C-index from 0.745 (95% CI, 0.723–0.767) to 0.747 (95% CI, 0.725–0.769), while adding plaque count changed it to 0.748 (95% CI, 0.726– 0.770). However, minor yet significant improvements were observed in the integrated discrimination improvement (IDI): 0.002 (95%CI, 2×10^-4^–0.005) for plaque presence and 0.002 (95%CI, 2×10^-5^–0.006) for plaque count. All the models demonstrated good calibration, indicating a strong alignment between observed outcomes and predicted risk estimates (**Supplementary Figure 12**).

To assess whether plaque phenotyping would improve the reclassification of individuals when complementing established clinical risk prediction models, we calculated the PCE risk scores for UKB participants eligible for assessment according to ACC/AHA guidelines^6^. Due to the PCE’s tendency to overestimate risk in the UKB population, the model was recalibrated (**Supplementary Figure 5**). Incorporating plaque presence and count into the PCE demonstrated significant reclassification improvement, with a categorical NRI of 0.034 (95% CI, 0.006–0.062) and 0.04 (95% CI, 0.010–0.070), respectively, at the threshold of 7.5% 10-year cardiovascular risk, which defines intermediate risk and justifies preventive initiation of statin therapy according to current guidelines. Overall, adding plaque presence correctly reclassified 17 out of 318 patients who developed MACE into a higher risk category. Including plaque count correctly reclassified 20 patients into a higher risk category (**Table 3**). In both cases–plaque presence and plaque count–four patients were incorrectly reclassified into a lower risk category.

**Table 3.**
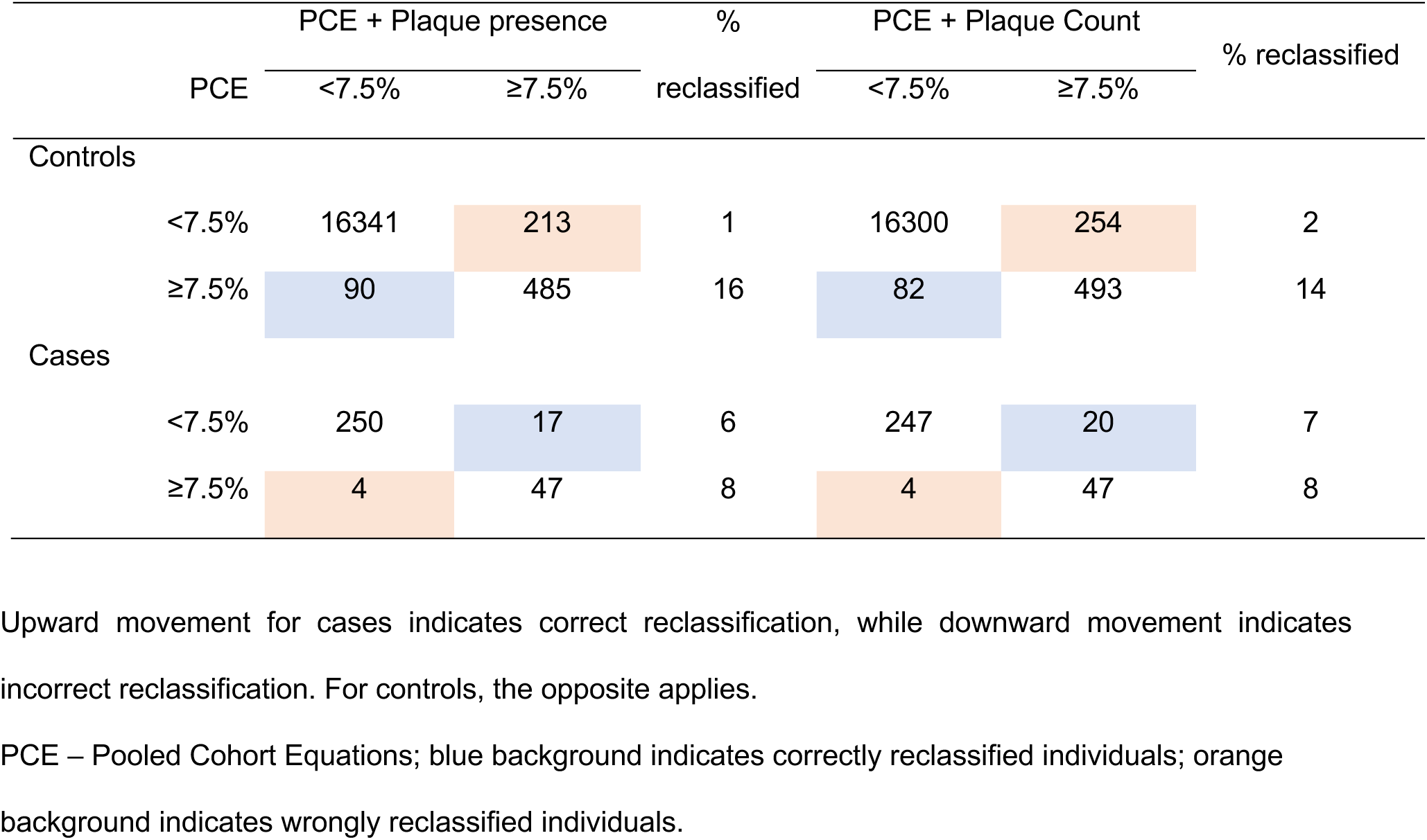
Reclassification table showing the distribution of individuals who went on to develop or not major adverse cardiovascular events into two risk groups based on the recommended 7.5% threshold from PCE risk assessment, both with and without incorporating plaque information.

### Genome-wide association study of carotid atherosclerotic plaque

As the next step, we investigated the genetic architecture of carotid atherosclerosis, defined by plaque presence. To detect single nucleotide variants (SNVs) associated with the presence of carotid atherosclerotic plaque, we conducted a genome-wide association study (GWAS) and subsequently meta-analyzed our data with the largest available GWAS for carotid plaque from the Cohorts for Heart and Aging Research in Genomic Epidemiology (CHARGE) consortium^74^. After quality control and excluding individuals without genetic data, the UKB GWAS included 18,203 White British individuals, comprising 8,250 cases with carotid plaque and 9,953 controls. The pooled sample from the UKB and CHARGE cohorts included 66,637 individuals (29,790 cases; 36,847 controls). We identified seven independent genomic loci significantly associated with the presence of carotid plaque, two of which were novel. Five of the loci (mapped to the genes *EDNRA, LINC02577, CDKN2B-AS1, CFDP1, LDLR*) replicated known associations, as the lead SNVs were in high linkage disequilibrium (r^2^ > 0.9) with SNVs previously reported to be associated with carotid plaque presence in the CHARGE study (**Figure 5**, **Supplementary Table 8**). The sixth locus included the *LPA* gene, which encodes lipoprotein(a) (Lp(a)) and is a known locus for atherosclerotic cardiovascular disease^75–79^. The lead variant at this locus (rs56393506), associated with higher odds for an atherosclerotic plaque (OR for T allele: 1.12, 95%CI: 1.07–1.16), is an intronic variant in the *LPA* gene that has been previously strongly associated with higher Lp(a) levels^80^. The lead SNV at the seventh locus is located in a non-coding region (rs1893250, OR for A allele: 0.93, 95%CI: 0.90–0.95) and was previously associated with angina pectoris^81^. Heterogeneity quantified by Cochran’s Q statistic was not significant for any of the genome-wide significant variants (**Supplementary Table 8**).

**Figure 5.**
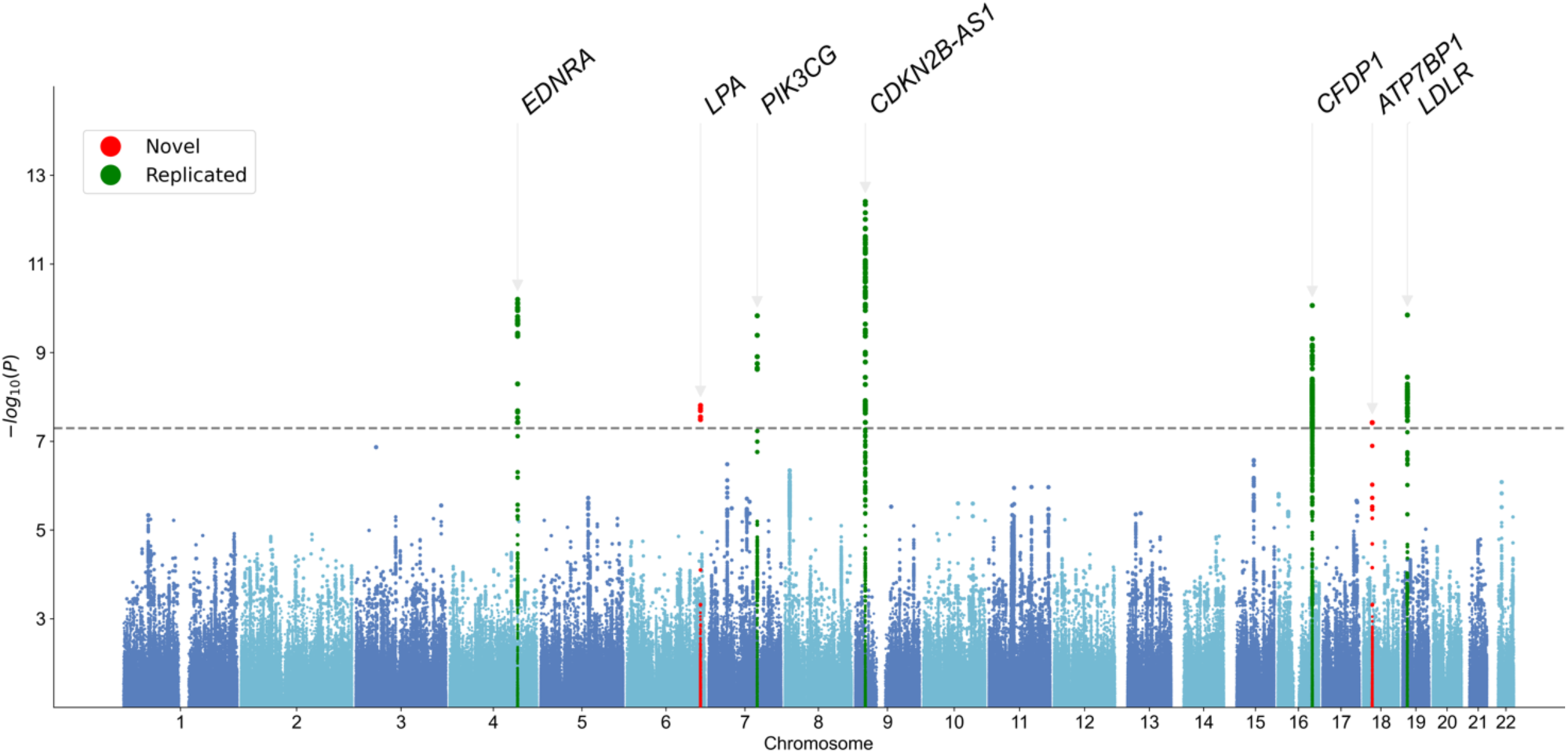
Manhattan plot of the GWAS meta-analysis for carotid plaque presence (29,790 cases; 36,847 controls). Loci highlighted in red point to novel significant associations for carotid plaque whereas loci highlighted in green represent validation of previously described associations.

### Mendelian randomization analyses

Finally, we performed Mendelian Randomization (MR) to explore whether genetically proxied risk factors and biomarkers of CVD are associated with carotid atherosclerotic plaque. We used the largest publicly available GWAS summary statistics to date for vascular risk factors to generate genetic instruments for the risk variables under study (**Supplementary Table 3**). The inverse-variance weighted (IVW) MR analyses revealed associations between higher genetically proxied SBP, diastolic blood pressure (DBP), LDL cholesterol, interleukin-6 (IL-6) signaling activity, and genetic predisposition to smoking initiation and type 2 diabetes (T2D) with the odds of carotid plaque presence (**Figure 6**). Additionally, higher genetically proxied HDL cholesterol was associated with lower odds of carotid plaque (**Figure 6**). The results for the significant IVW associations were generally highly consistent in sensitivity analyses, including MR-Egger regression and the Weighted Median estimator (**Supplementary Table 9**).

**Figure 6.**
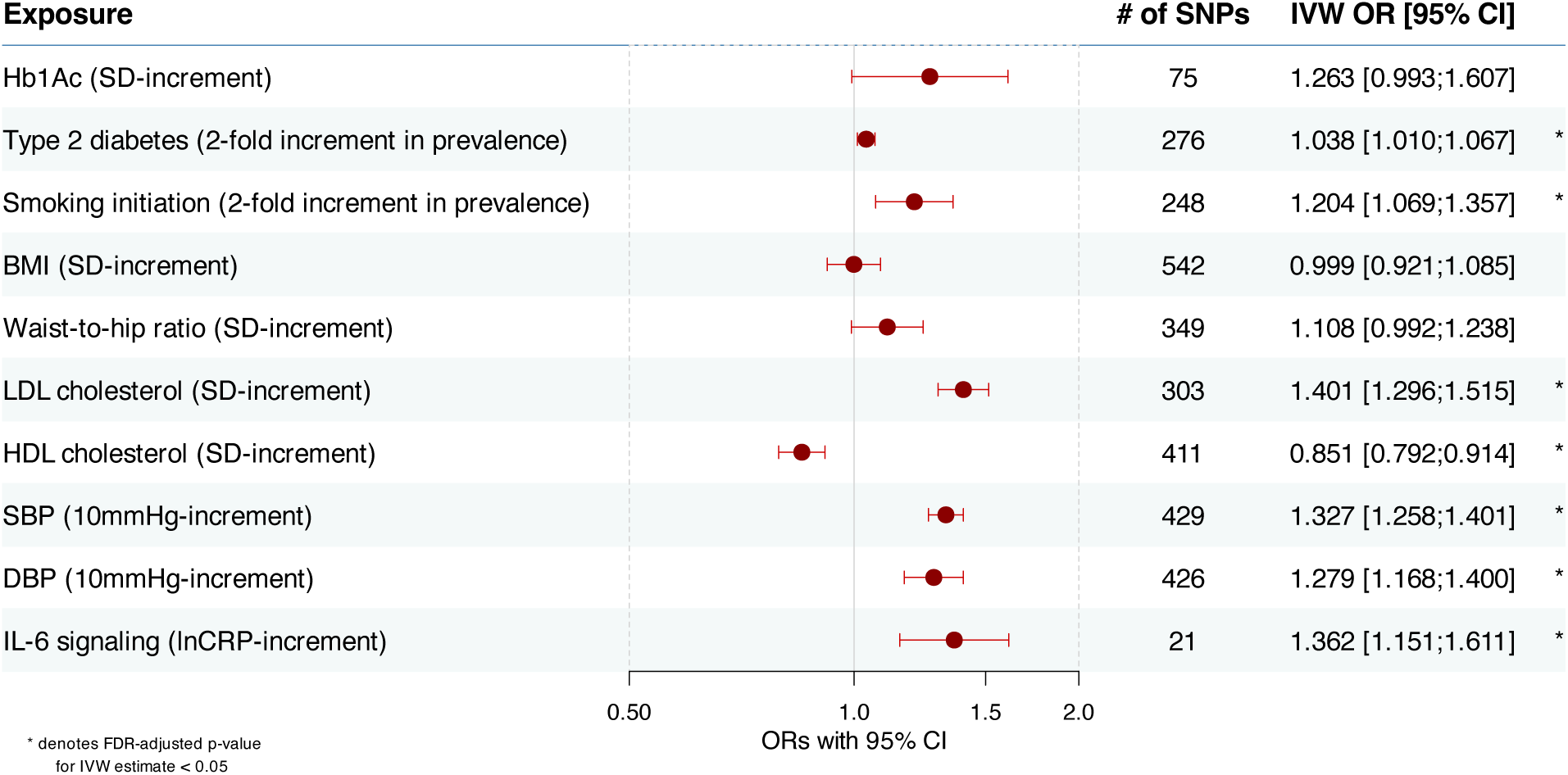
Forest plot for Mendelian randomization results. The results are presented as odds ratios (OR) and 95% confidence intervals (CI) derived from random-effects inverse-variance weighted Mendelian randomization analyses. Two-fold increments in prevalence for binary exposures (type 2 diabetes and smoking initiation) were derived by multiplying the IVW betas and corresponding confidence intervals by 0.693, as described by Burgess and Labrecque^97^. HbA1c – Glycated hemoglobin; BMI – Body Mass Index; LDL – Low-Density Lipoprotein Cholesterol; HDL – High-Density Lipoprotein Cholesterol; SBP – Systolic Blood Pressure; DBP – Diastolic Blood Pressure; IL-6 – Interleukin-6.

## Discussion

In this study, we developed a computer vision model that accurately detects atherosclerotic plaques in carotid ultrasound images and applied it to a population-based cohort of 19,499 participants from the UKB. The model demonstrated strong performance in detecting carotid plaques and classifying plaque-positive carotid ultrasound images, achieving approximately 90% in accuracy, sensitivity, and specificity. Following fine-tuning, the model also achieved high performance in the external BiDirect cohort, which followed a different scanning protocol with another ultrasound device. Consistent with previous studies in comparable demographics (mean age 64.6±7.6 years; 53% female)^14,82^, our model identified at least one carotid plaque in 45% of the UKB participants. The presence of plaques was associated with conventional vascular risk factors and was predictive of future adverse cardiovascular events over a follow-up period of up to 7 years. Importantly, both plaque presence and count led to improved risk reclassification for future adverse cardiovascular events beyond the established PCE risk assessment tool. Leveraging the phenotypic depth of the UKB, we conducted the largest genomic analysis of carotid atherosclerosis to date, identifying two novel loci and risk pathways, including ones targeted by emerging cardiovascular therapeutics, such as Lp(a) and IL-6 signaling. The developed model enriches the UK Biobank with important information on carotid plaque presence and count, and offers a promising solution for plaque phenotyping in large-scale cohorts.

While atherosclerosis can develop long before clinical symptoms appear^83^, modern risk assessment tools do not consider subclinical atherosclerotic pathology. Many studies have highlighted the potential of integrating imaging biomarkers of subclinical atherosclerosis into conventional risk assessment tools^28^. CAC assessed through CT is a well-established predictor of CVD risk^84^. However, CAC has limitations, including insensitivity to early stages of atherosclerosis and exposure to radiation^19,20,85^. In contrast, carotid ultrasound is an inexpensive, well-tolerated, radiation-free tool capable of detecting early-stage atherosclerosis^14,86^. Our results, based on a population of almost 20,000 individuals, suggest that both the presence and count of plaques are strongly associated with the risk of future acute CVD events. Incorporating plaque information into Cox regression models led to significant reclassification improvements, which were robust across the full cohort, individuals without a history of CVD, and statin-naive participants. The reclassification metrics in our study indicate that adding plaque information to conventional risk factors and directly incorporating it into the PCE has the potential to improve patient stratification. Specifically, incorporating plaque count reassigned 6.3% (20 out of 318) of individuals who went on to develop cardiovascular events from a low to a higher risk category, making them eligible for preventive statin therapy.

Despite growing evidence that carotid ultrasound-derived plaque phenotypes—such as total plaque area and vulnerability features—are independently associated with future CVD events ^26,87,88^, their use in clinical practice remains underutilized. The primary challenges for wider adoption include the time required for assessment and reliance on operator skill^23^. However, an efficient artificial intelligence model could significantly streamline these labor-intensive tasks. The model developed in this study demonstrates high performance in identifying individuals with carotid atherosclerosis and localizing plaques from a single image of the carotid bifurcation. Further model advancements could not only address the issue of labor intensity but also enable the extraction of more detailed plaque features, thereby improving the predictive performance of carotid ultrasound.

Importantly, our model enhances the phenotypic depth of the UKB to include subclinical atherosclerosis, facilitating integration with the unique resources available in this population, such as multi-omics and other imaging data^89^. Moving in this direction, we leveraged the available genomic data to perform the largest exploration to date of the genetic architecture of carotid plaque. We replicated five genomic loci previously associated with atherosclerosis endophenotypes^74^ and also found two new loci related to clinical cardiovascular outcomes. Furthermore, downstream MR confirmed the effect of genetic predisposition to known vascular risk factors, such as smoking, high blood pressure, and LDL cholesterol, on carotid plaque presence. Importantly, the GWAS and MR results showed that genetic variation leading to elevated Lp(a) levels and higher IL-6 signaling activity is associated with higher odds of carotid plaque. Both pathways are believed to play key roles in atheroprogression and are the targets of investigational drugs in advanced clinical development^75,76,90–92^. These results suggest that drugs targeting these pathways could be promising, particularly in the preclinical stages of atherosclerosis. Integrating carotid plaque phenotypes with additional omics layers may provide further insights into novel drug targets for atherosclerotic CVD.

### Limitations

Our study has several limitations. First, during model development, the low quality of some images necessitated contrast enhancement and noise reduction techniques. These adjustments may have introduced bias, especially in cases where high noise levels complicated plaque detection. However, our model achieved high classification and detection metrics, which could potentially be improved further by annotating more images. Moreover, the consistency of predicted carotid plaque prevalence with previously reported plaque prevalence in similar demographics, along with the associations of predicted plaque presence with known risk factors and future CVD risk, supports the model’s reliability^82^. Second, the UKB is a cohort of healthy volunteers with a lower incidence rate of CVD than the general population, particularly within its imaging subsample (**Supplementary Table 6**)^93^. This discrepancy contributes to an overestimation of CVD risk calculated by clinical risk models, such as the PCE tool. To improve the reliability of our reclassification evaluation, we recalibrated the model using data from the study population to better align predicted risk with observed outcomes. Due to the low prevalence of risk factors and events, PCE-derived absolute risk estimates remain notably low. It is worth noting that deriving categorical NRI metrics comparable to those from other studies with different incidence rates may be problematic^94^. We addressed this issue by calculating continuous NRI estimates using bootstrap estimates, which are threshold-independent and less sensitive to event rates^95^. Third, the carotid ultrasound examination took place 2 to 15 years after the baseline visit and the assessment of cardiovascular risk factors. This gap introduces bias into the effect estimates. To account for changes in baseline risk factors over this period, we used, wherever available, data from the primary care records of participants collected at the closest date to the ultrasound exam. Fourth, our study has a shorter follow-up period of 7 years compared to most clinical risk assessment tools, including the PCE, which typically calculate 10-year risk estimates for CVD. Fifth, we observed significant heterogeneity in the results of IVW MR analyses for several risk factors (**Supplementary Table 9**), which could indicate the presence of pleiotropy. To explore whether the derived estimates could be biased by directional pleiotropy, we conducted several sensitivity analyses to test the robustness of the estimates against different MR assumptions. Sixth, there was some population overlap between the exposure and outcome GWAS datasets used in our MR analyses, which could introduce weak instrument bias into the derived effect estimates. We addressed this concern by using the largest available summary statistics for the exposure data. Given that the effective population overlap was less than 5% for all exposure-outcome pairs, we estimated any bias in the effect estimates to be under 5%^96^. Seventh, all GWAS used for the MR analyses were conducted in participants of European ancestry; therefore, the estimates primarily generalize to European populations and may not be applicable to other ancestries. Eighth, among some of the CHARGE cohorts included in the GWAS meta-analysis, carotid plaque was defined using criteria different from the Mannheim consensus used in this study. This variation may introduce phenotypic heterogeneity that could affect the meta-analysis results. However, all definitions employed are based on standard clinical criteria and reflect the presence of atherosclerotic arterial pathology. Another limitation lies in differences between the UKB and external validation cohort. UKB is population-based, whereas BiDirect includes a relatively high proportion of participants with cardiovascular disease (15%; see **Supplementary Table 7**) and a high prevalence of clinical depression (45%). The cohorts also used different ultrasound protocols, devices, and image resolutions, necessitating model fine-tuning. Additionally, the BiDirect sample was relatively small (N=2,105) compared with the UKB (N=19,499), which limits the precision of the prevalence estimates. Lastly, this study analyzed samples from the UK and Germany, which predominantly consisted of White ancestry volunteers, who are healthier than the general population. Therefore, replicating these findings in more diverse populations and real-world settings is crucial for improved generalizability and informed decision-making.

### Conclusions

We have successfully developed, implemented, and externally validated a deep learning model for early atherosclerotic plaque detection in carotid ultrasound imaging. Our model sets the stage for automating subclinical carotid plaque assessment in the context of either population screening for asymptomatic individuals or large-scale plaque phenotyping in population-based cohorts. Our results highlight the potential of large-scale carotid plaque assessment for refining cardiovascular risk prediction and revealing relevant biological mechanisms for atherosclerosis.

## Data Availability

The UKB provides an accessible research resource, available to researchers upon submitting a research proposal at https://www.ukbiobank.ac.uk. The GWAS data on UKB and the GWAS meta-analysis results obtained in this study are available in the GWAS Catalog (https://ftp.ebi.ac.uk/pub/databases/gwas/summary_statistics/GCST90454001-GCST90455000/) under accession numbers GCST90454354 and GCST90454353, respectively. Additionally, the carotid plaque phenotypes derived from the developed model will be returned to UKB for use in future studies. The list of GWAS used for MR analyses are presented in Supplementary Table 3. Individual-level data of the BiDirect cohort can be obtained by researchers from KB (https://medizin.uni-muenster.de/en/epi) following a written transfer agreement.
The code used for this study will be available on GitHub upon publication of this manuscript.

## Acknowledgements

UKB data were accessed through applications 36993, 7089, and 151281. The graphical abstract was created using BioRender.

