## Supplementary Figures for "Automated Deep Learning-Based Detection of Early Atherosclerotic Plaques in Carotid Ultrasound Imaging"

**Supplementary Figure 1.** Flowchart of the study participants and corresponding carotid ultrasound images.

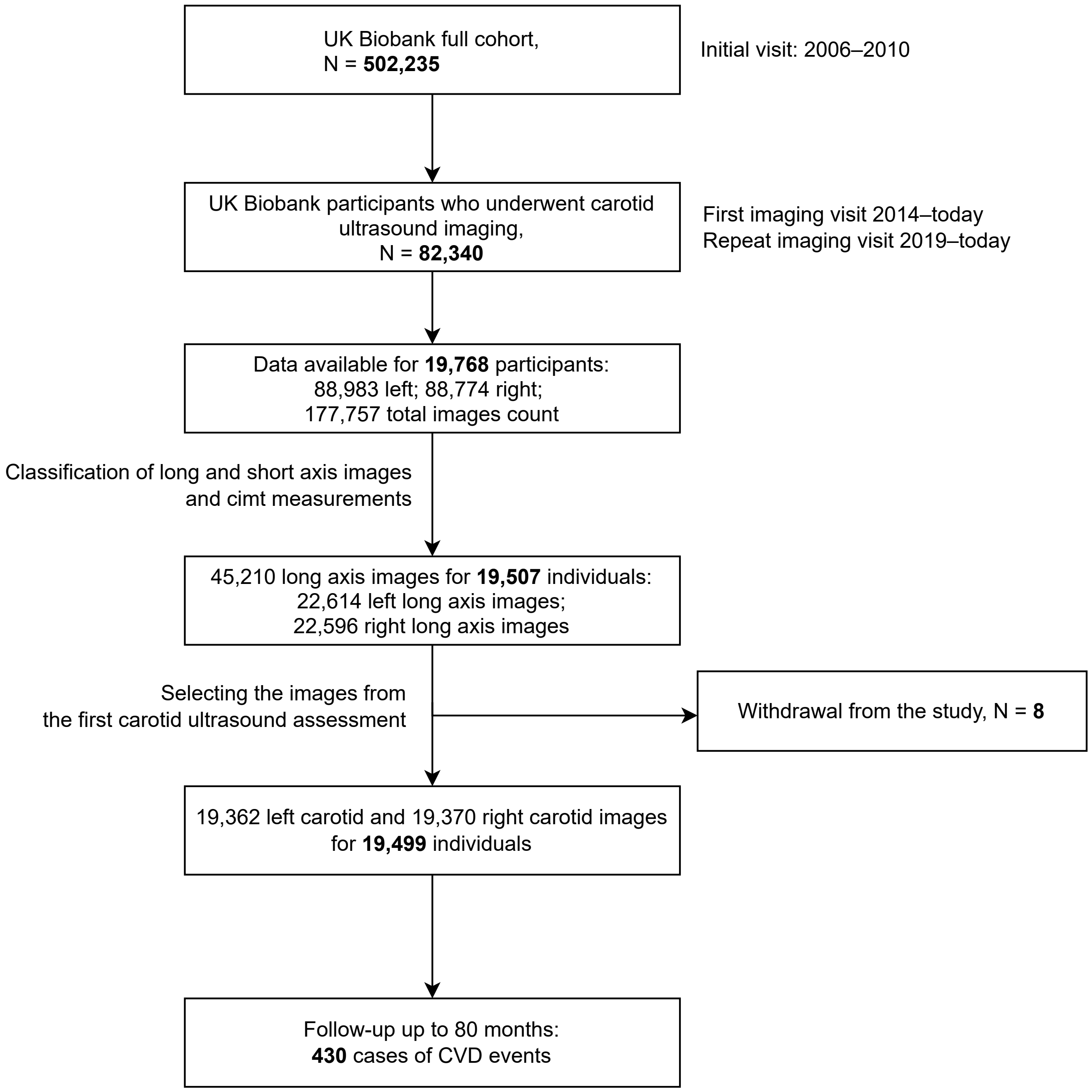

**Supplementary Figure 2.** To categorize all the images according to their type, we wrote a Python script that counts the number of pixels in specific locations of the ultrasound machine picture frame, tracking the angle and type of examination. The obtained results were manually validated. The classified long axis images were cropped to a size of 480x448 to keep only the ultrasound image while maintaining the original resolution. In order to enhance contrast and decrease noise in the images, we applied two functions from the OpenCV library: median blur filtering (*ksize=5*) and Contrast Limited Adaptive Histogram Equalization (*clipLimit=2.0, tileGridSize=(8,8)*) respectively.

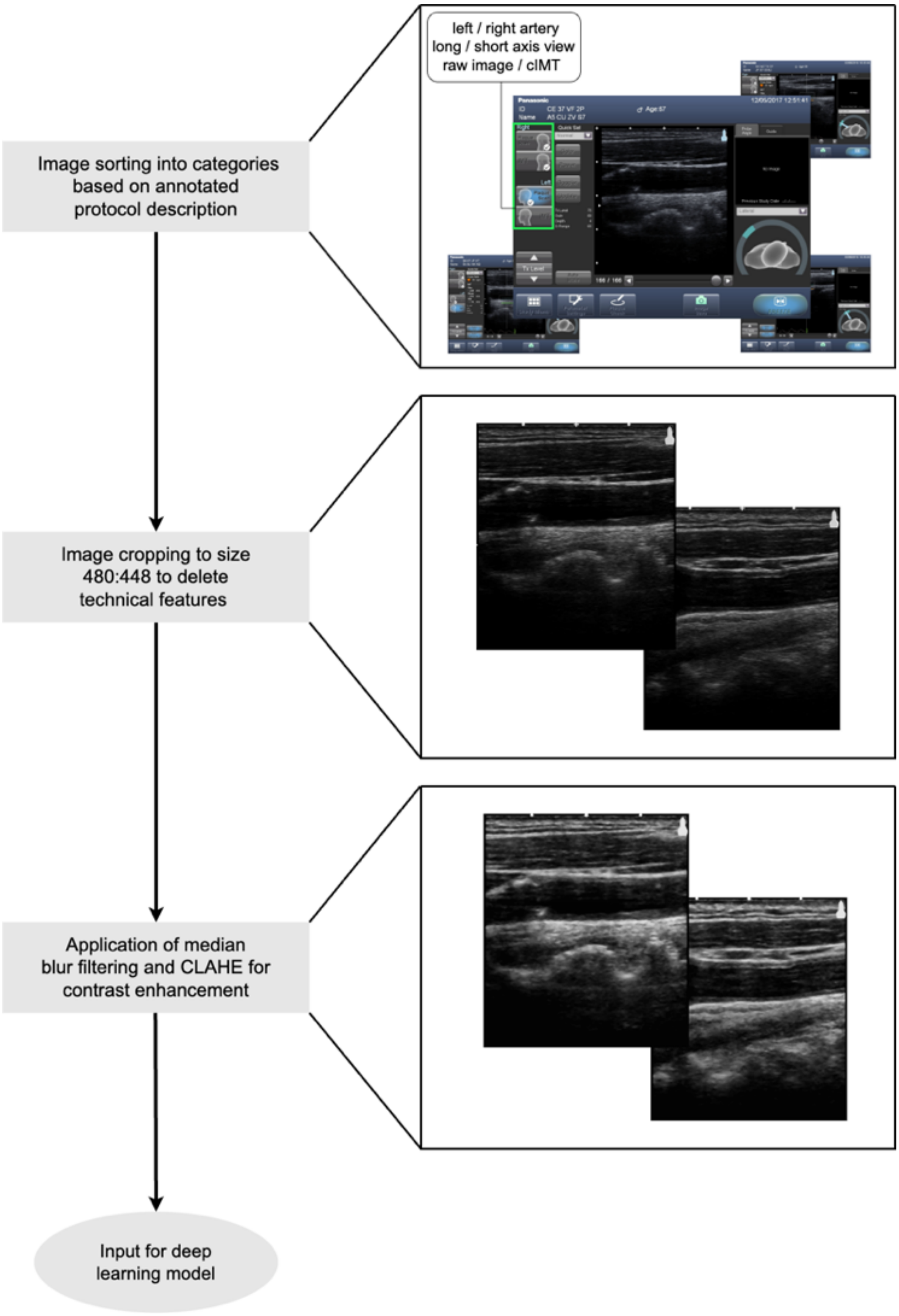

**Supplementary Figure 3.** Model segmentation output for image selection in the BiDirect cohort. **A**. Comparison of manual annotations with lumen segmentation model predictions. **B.** Predicted binary segmentation masks before and after post-processing.

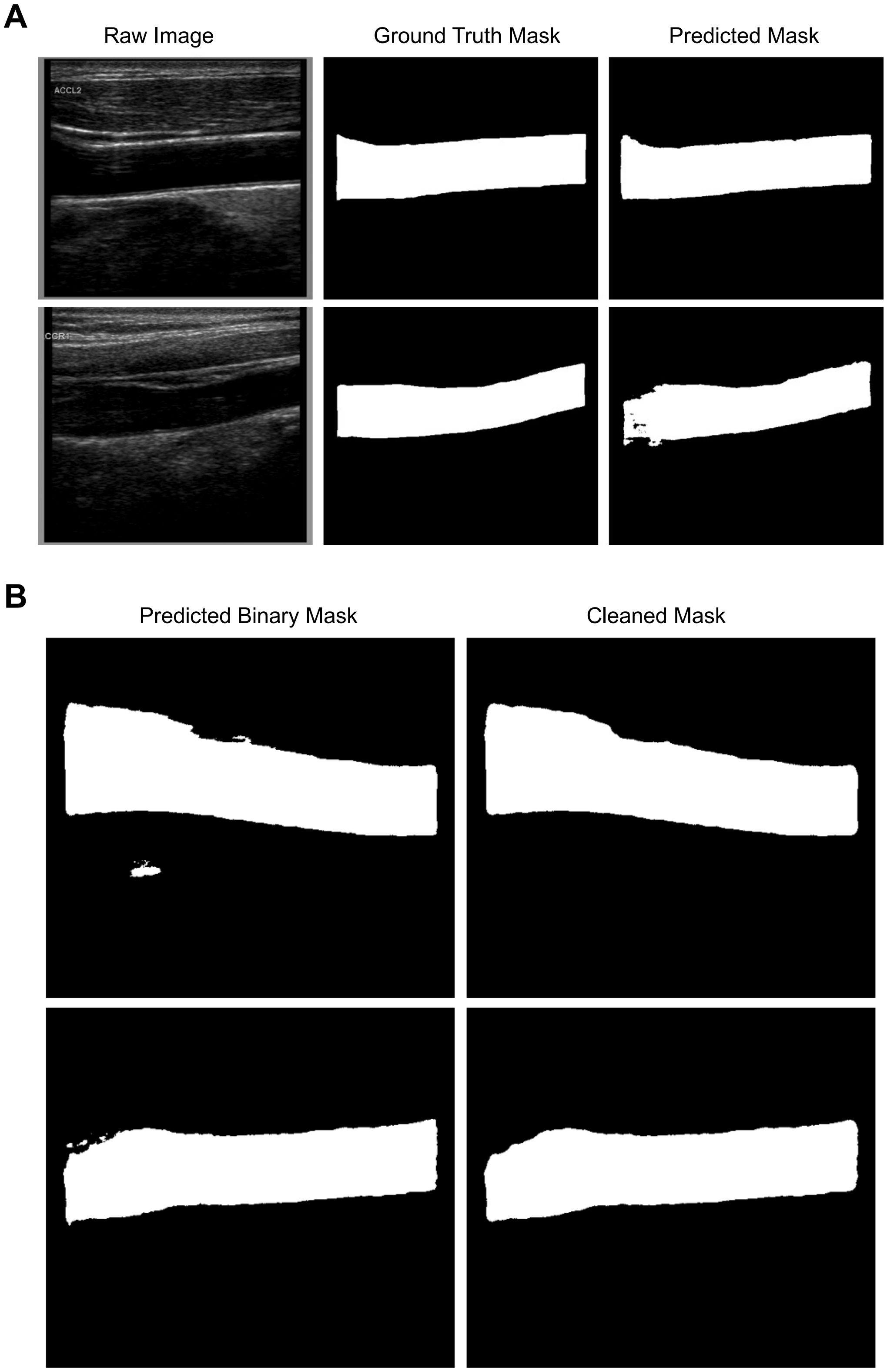

**Supplementary Figure 4.** Results of the 5-fold cross-validation performed on a combined training and validation dataset. Model parameters were set as specified in the Methods section.

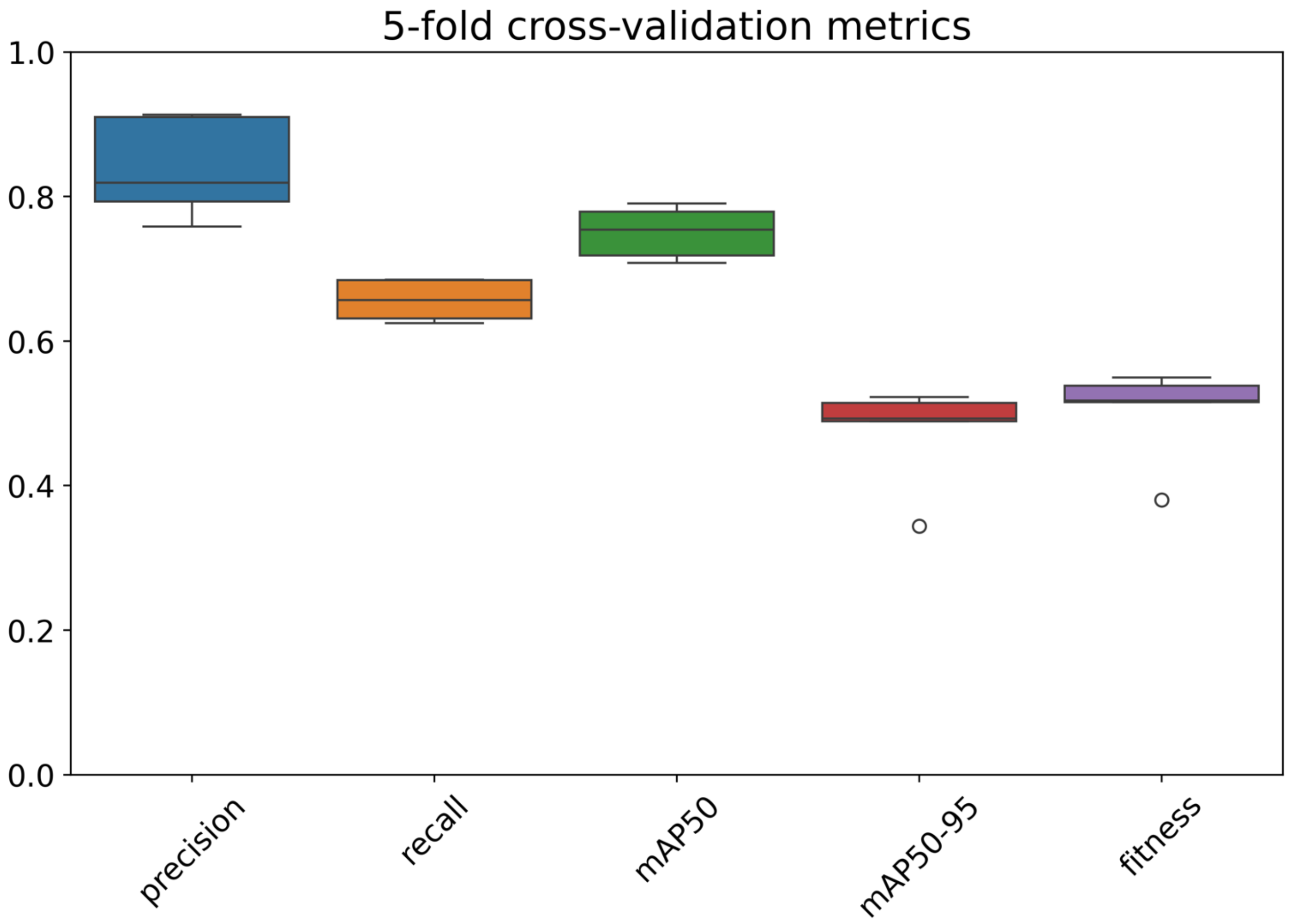

**Supplementary Figure 5.** Calibration plots for the Pooled Cohort Equations (PCE) applied to the UK Biobank cohort before and after recalibration and incorporating plaque information

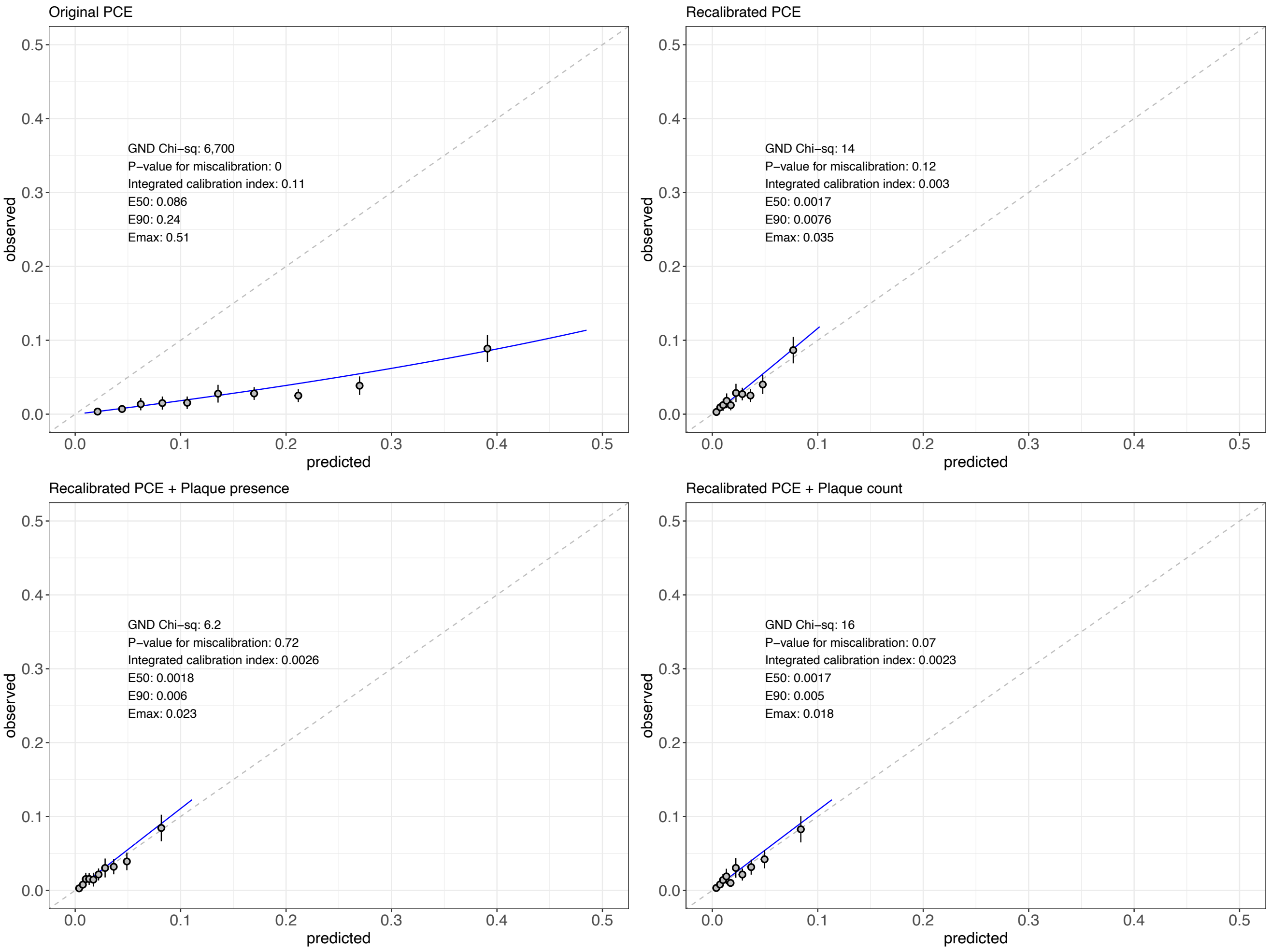

**Supplementary Figure 6.** Examples of the model's predictions in the UK Biobank.

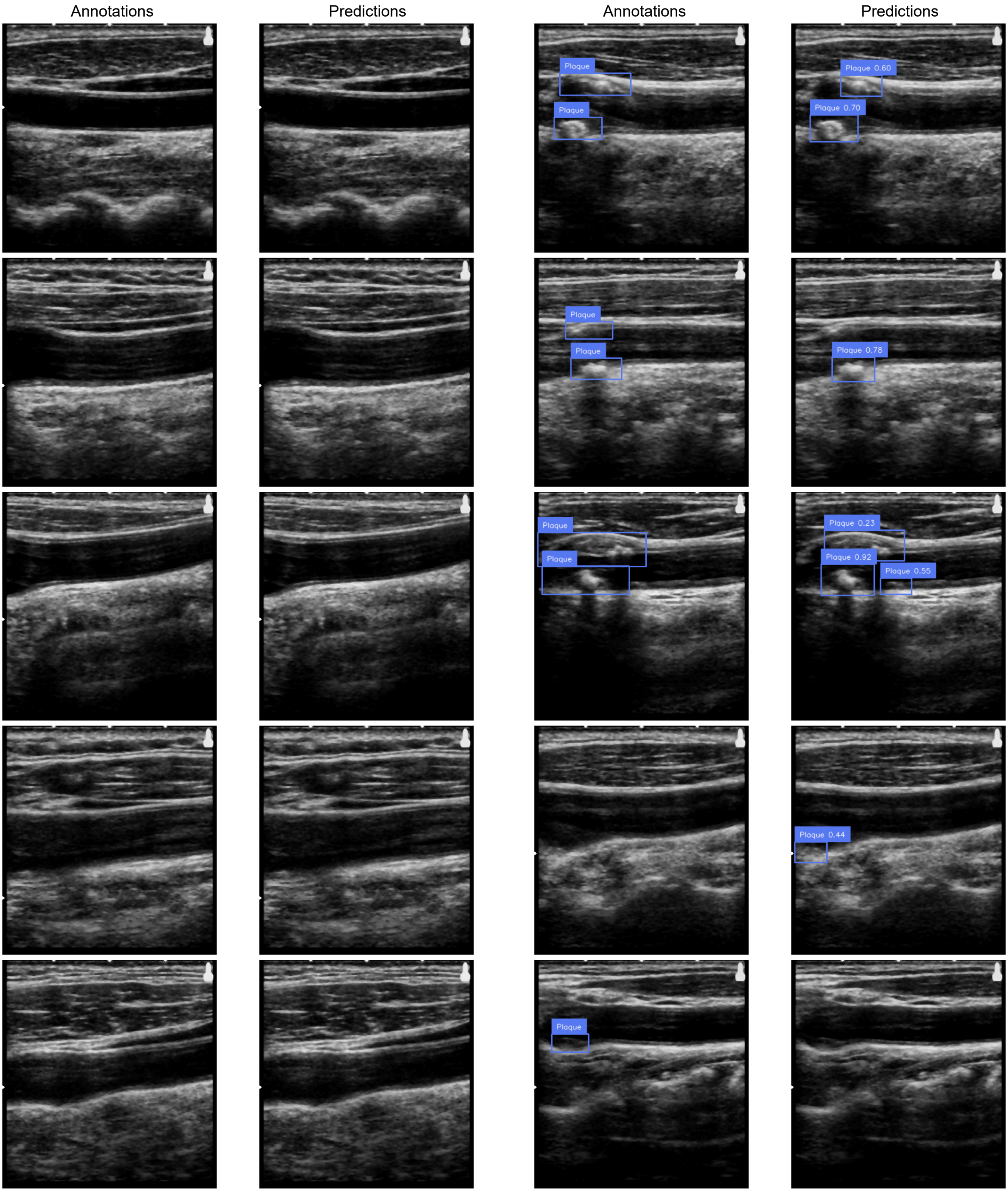

**Supplementary Figure 7.** Confusion matrix for plaque detection in the UK Biobank test set (n = 103 images). Every count represents a single plaque as a distinct object. FP – False Positive; FN – False negative.

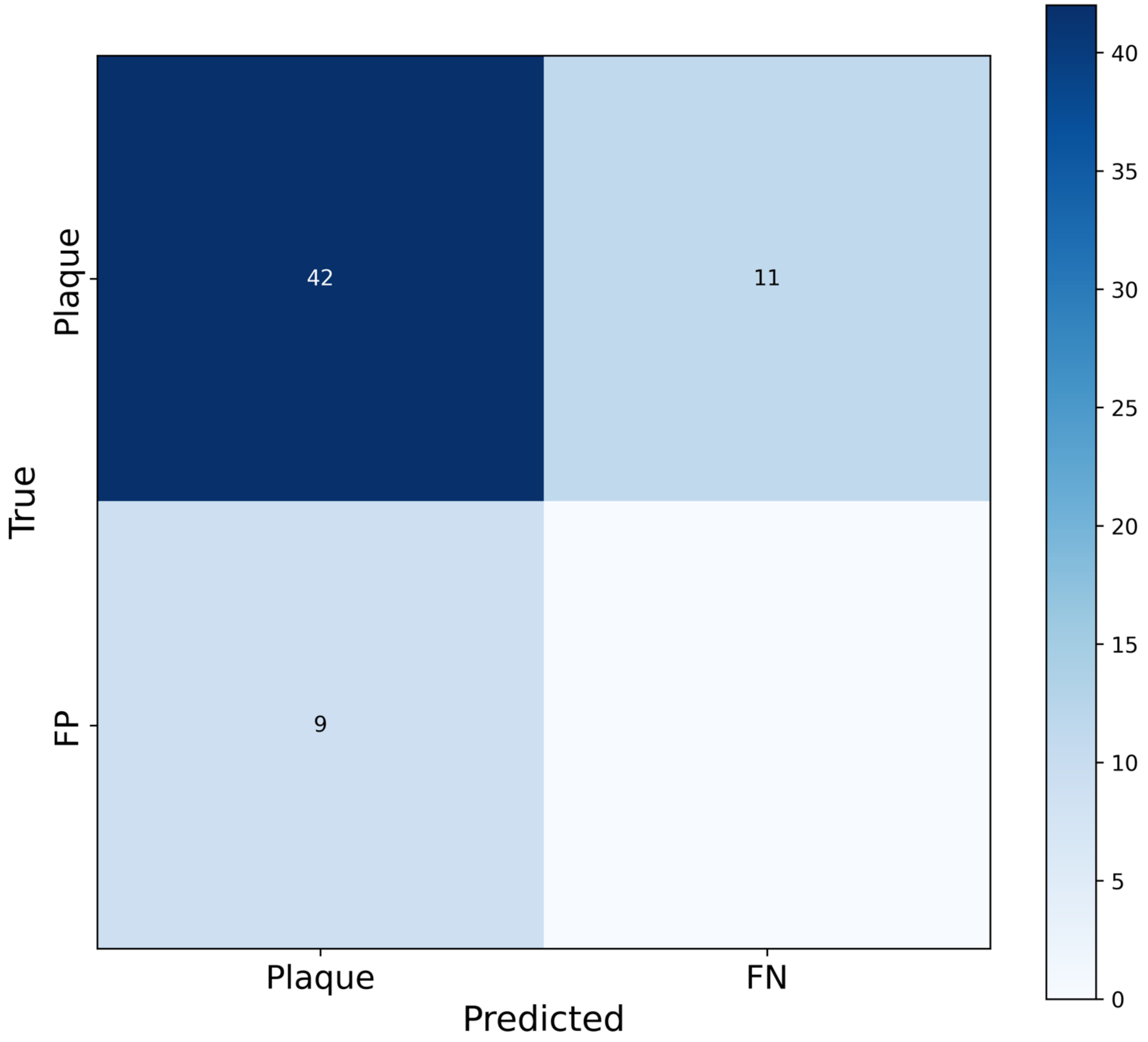

**Supplementary Figure 8.** Comparisons of three carotid intima-media thickness (IMT) measurements between individuals with model-predicted plaque and those without in the UK Biobank. **A)** Averaged measurements of both left and right carotid arteries; **B)** Left carotid artery; **C)** Right carotid artery

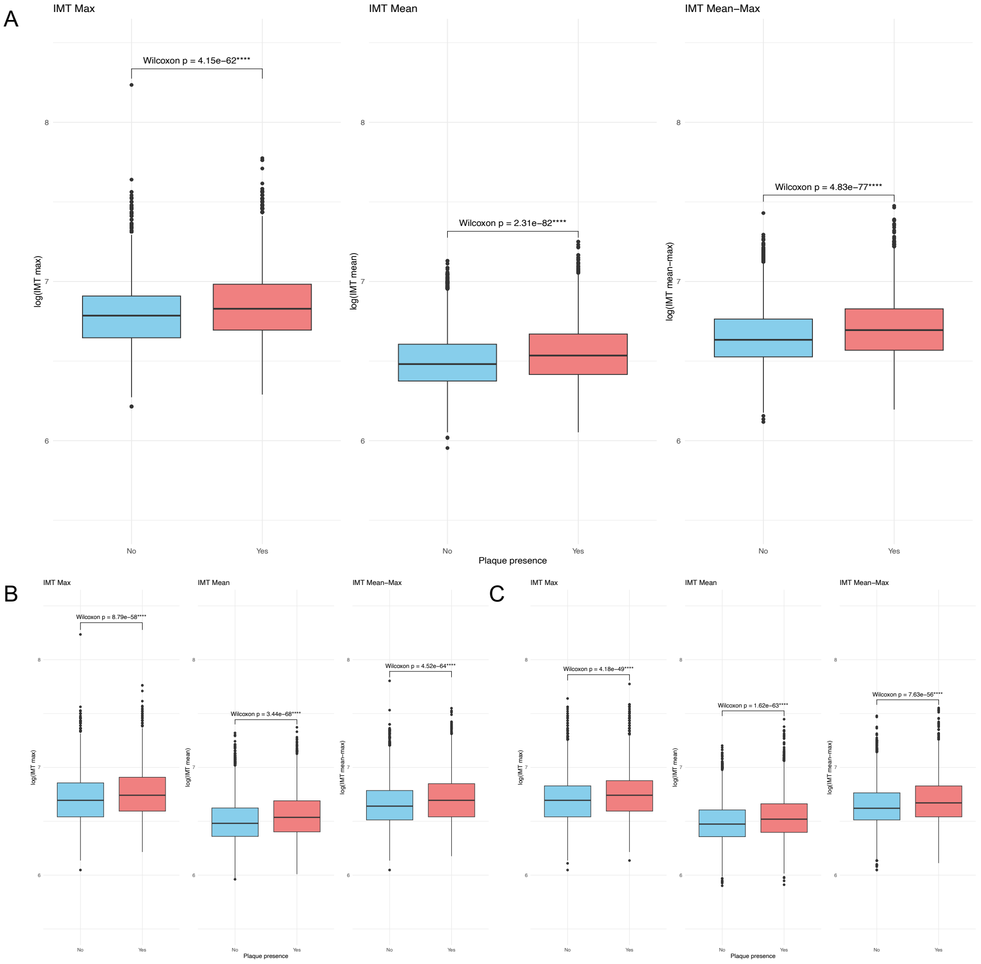

**Supplementary Figure 9.** Confusion matrix for plaque detection in the BiDirect test set (n = 120 images). Every count represents a single plaque as a distinct object. FP – False Positive; FN – False negative.

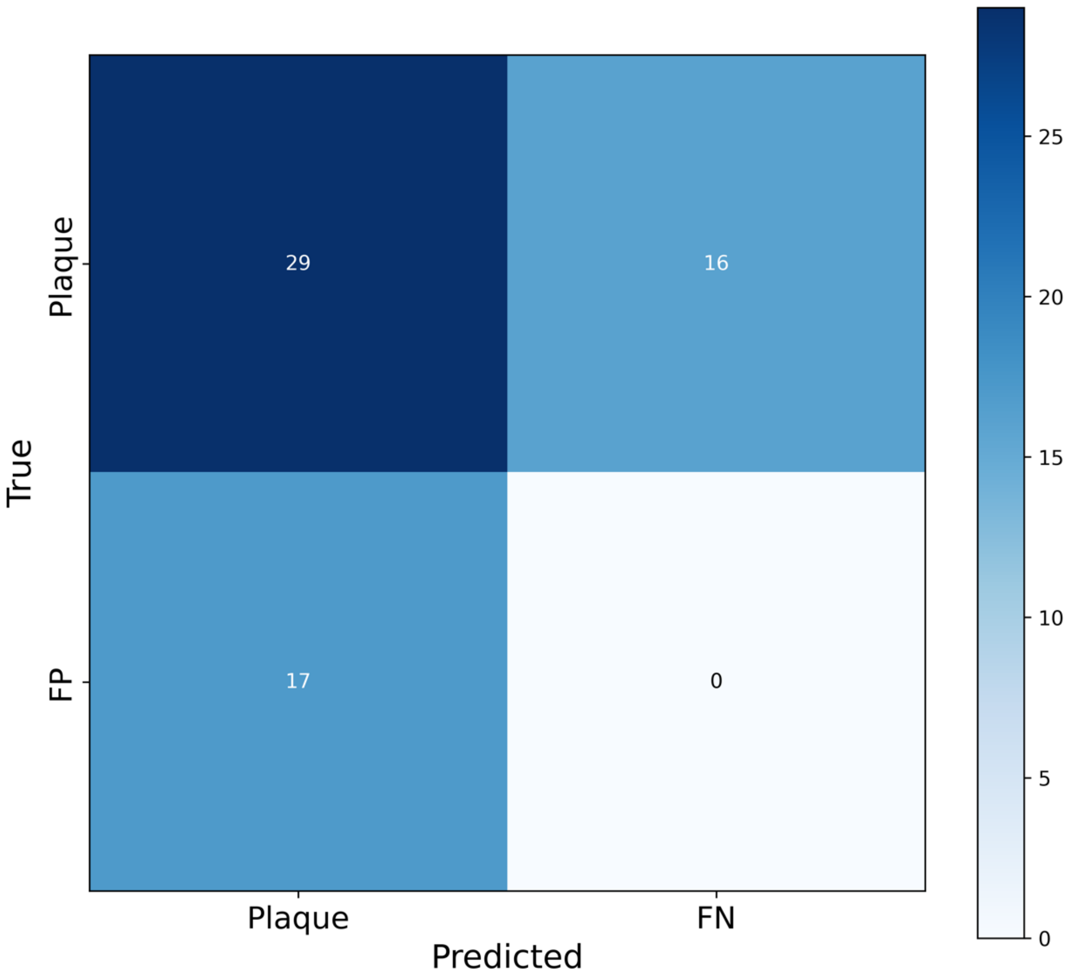

**Supplementary Figure 10.** Comparisons of mean carotid intima-media thickness (IMT) measurements between individuals with model-predicted plaque and those without in the BiDirect cohort. **A)** Left carotid artery; **B)** Right carotid artery

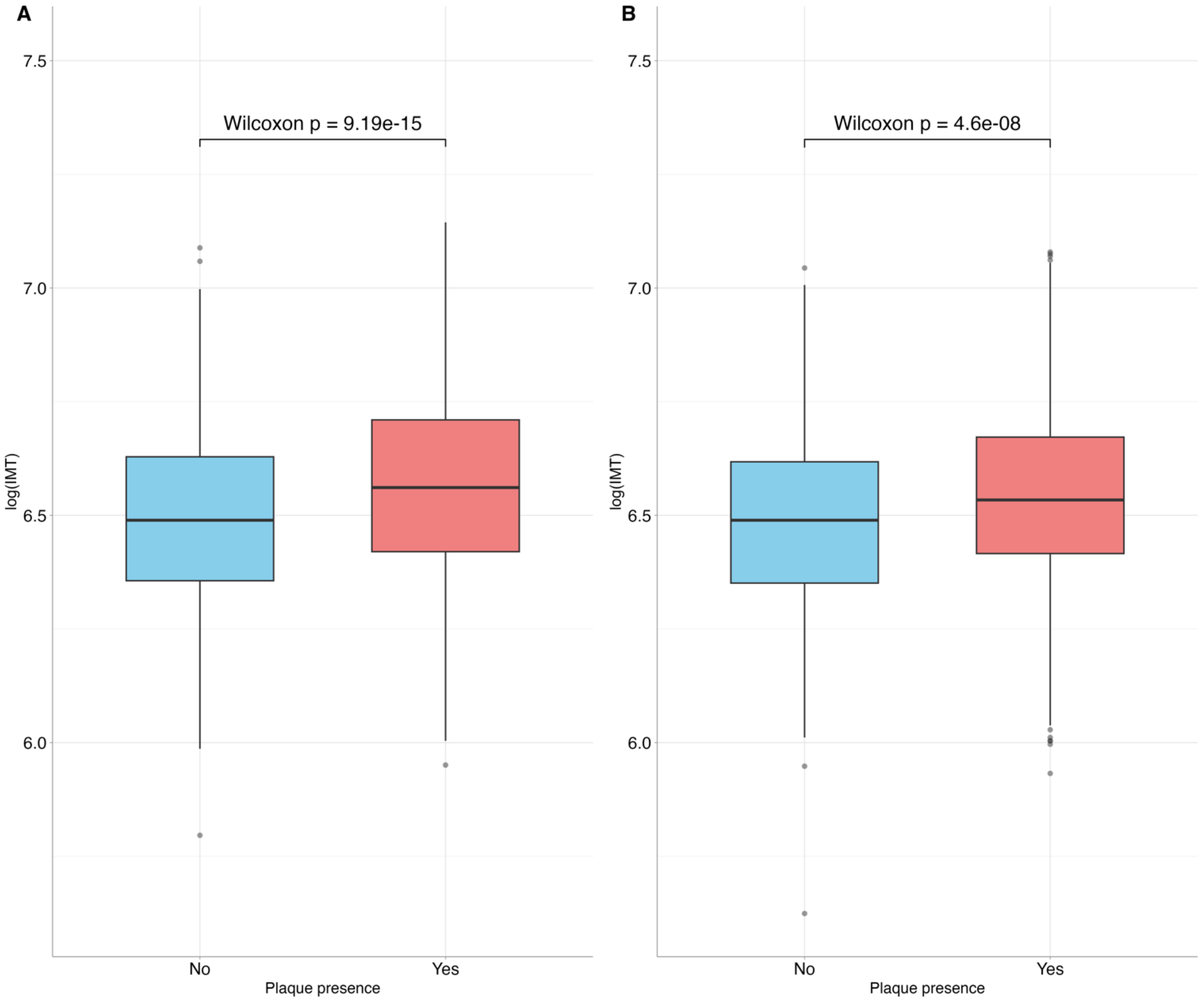

**Supplementary Figure 11.** Hazard ratios (HRs) from Cox models fitted on different subsamples for **A)** Major Adverse Cardiovascular Events (MACE) and its individual components: **B)** Myocardial infarction; **C)** Stroke. HR estimates for plaque presence and plaque number (one plaque, two or more plaques) were obtained using two different models for each subsample. All plaque-related variables were encoded as categorical.

A

| Sample | Cases | Controls | HRs (95% CI) |
|---|---|---|---|
| Full sample | 430 | 19069 | 1.42 (1.16 to 1.73) |
| | | | 1.30 (1.04 to 1.63) |
| | | | 1.62 (1.27 to 2.07) |
| Primary events | 335 | 17775 | 1.39 (1.11 to 1.73) |
| | | | 1.29 (1.008 to 1.66) |
| | | | 1.57 (1.18 to 2.07) |
| Primary events (without statins) | 234 | 14283 | 1.38 (1.07 to 1.80) |
| | | | 1.32 (0.99 to 1.77) |
| | | | 1.51 (1.07 to 2.14) |

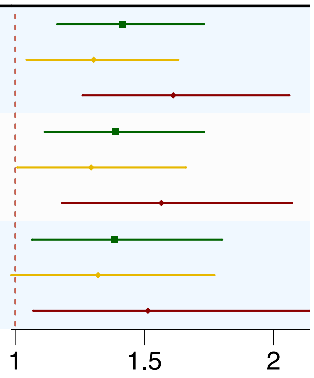


B

| Sample | Cases | Controls | HR (95% CI) |
|---|---|---|---|
| Full sample | 195 | 19304 | 1.46 (1.09 to 1.96) |
| | | | 1.39 (1.005 to 1.94) |
| | | | 1.58 (1.09 to 2.29) |
| Primary events | 155 | 17955 | 1.41 (1.02 to 1.95) |
| | | | 1.32 (0.92 to 1.91) |
| | | | 1.56 (1.03 to 2.37) |
| Primary events (without statins) | 106 | 14411 | 1.32 (0.89 to 1.95) |
| | | | 1.17 (0.75 to 1.83) |
| | | | 1.59 (0.97 to 2.62) |

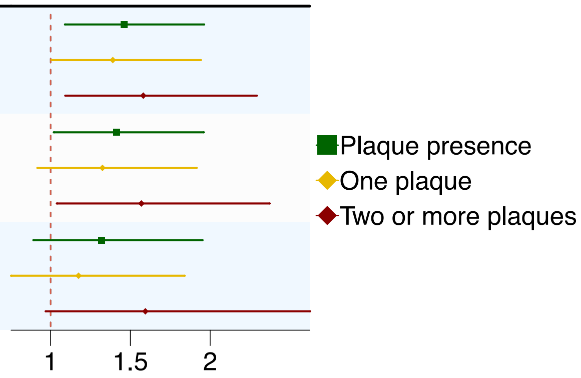


C

| Sample | Cases | Controls | HR (95% CI) |
|---|---|---|---|
| Full sample | 172 | 19327 | 1.30 (0.95 to 1.77) |
| | | | 1.20 (0.94 to 1.70) |
| | | | 1.47 (0.99 to 2.19) |
| Primary events | 136 | 17974 | 1.34 (0.95 to 1.89) |
| | | | 1.19 (0.84 to 1.70) |
| | | | 1.47 (0.98 to 2.19) |
| Primary events (without statins) | 103 | 14414 | 1.29 (0.87 to 1.92) |
| | | | 1.33 (0.86 to 2.05) |
| | | | 1.20 (0.68 to 2.09) |

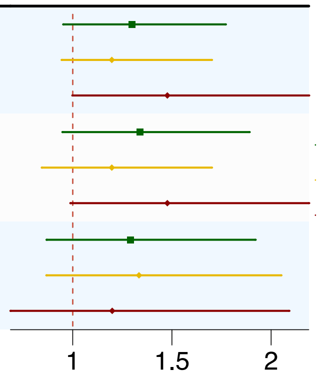

**Supplementary Figure 12.** Calibration plots for Cox models fitted on different subsamples **A)** Major Adverse Cardiovascular Events (MACE) and its individual components: **B)** Myocardial infarction; **C)** Stroke.

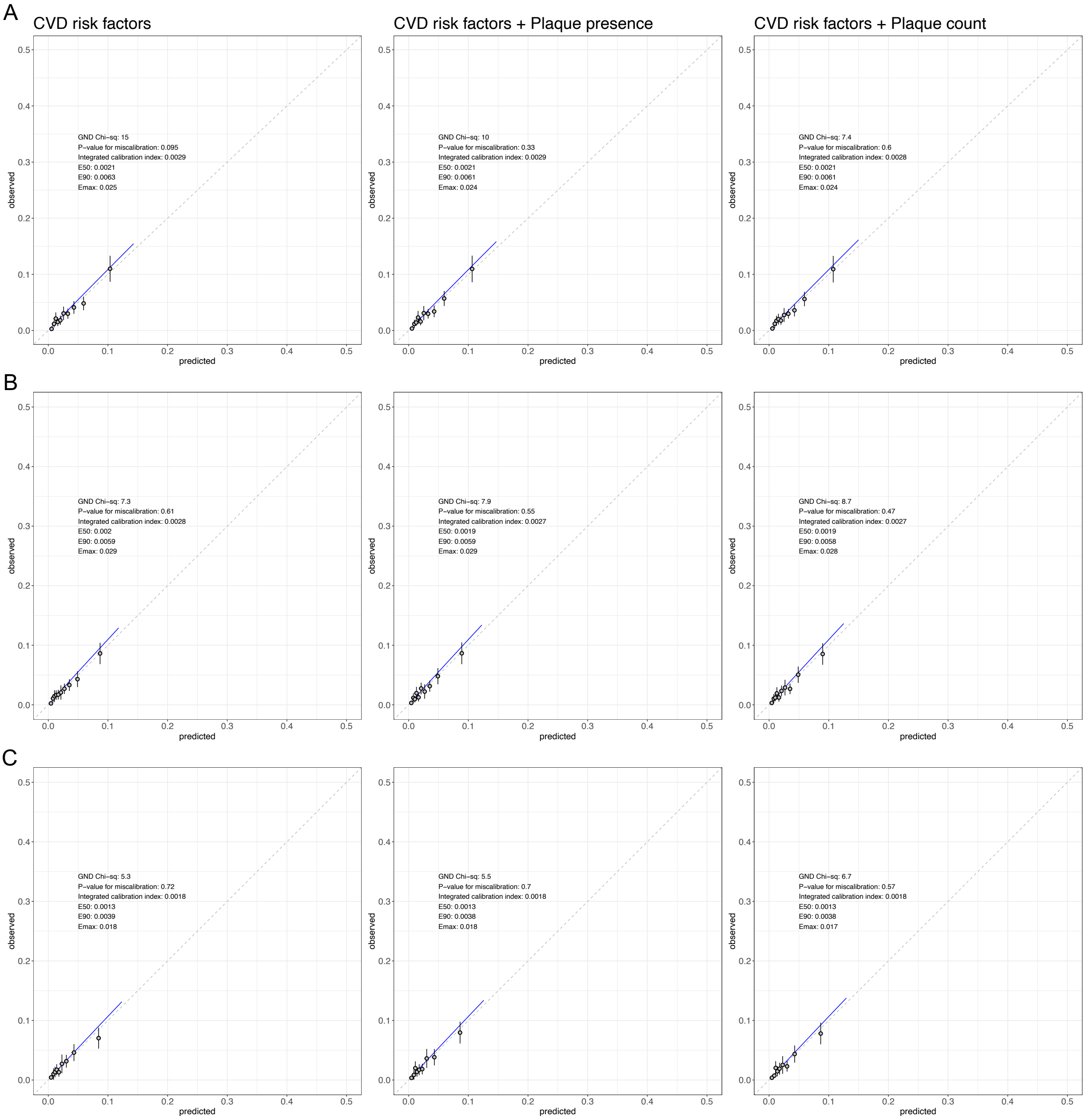
